# Diurnal variation in brain-derived tau and five other blood-based biomarkers for dementia and their association with cognitive performance

**DOI:** 10.64898/2026.06.12.26355520

**Authors:** Ciro della Monica, Samuel G. Stagg, Tegan Ward, Marta M. Pineda, Kiran K. G. Ravindran, Martina Del Giovane, Adam Hampshire, Amanda Heslegrave, Henrik Zetterberg, Simon S. Skene, Ramin Nilforooshan, Hana Hassanin, Victoria Revell, Derk-Jan Dijk

## Abstract

Blood-based biomarkers of dementia are a promising scalable tool for early diagnosis, tracking disease progression, and evaluating therapeutic efficacy. Utility of these biomarkers will not only be dependent on the reliability of their association with pathology but also contingent on their ability to track cognitive status. Previously, we demonstrated diurnal variation in several biomarkers (amyloid beta (Aβ) 42 and 40, 42/40 ratio, glial fibrillary acidic protein (GFAP), neurofilament light (NfL), and phosphorylated-Tau 217 (p-Tau217)) which has implications for their reliability. Here, we extend these observations to a larger cohort, include brain-derived tau (BD-Tau), which is assumed to be produced exclusively in the brain, and report endocrine measures of circadian rhythmicity. We not only assessed whether these biomarkers vary with time of day, but also whether they associate with daytime function and whether these associations vary with cognitive domain and number of repeated assessments.

Data collected in 20 PLWA (72.4±5.9 years, mean±SD) and 19 controls (68.9±9.8 years) were analysed. Participants completed 14 days of home monitoring and one laboratory assessment of sleep and daytime function: mood, daytime sleepiness, reaction time, immediate and delayed memory recall, everyday memory errors. During the 27-hour residential laboratory session, 3-hourly blood samples were collected and analysed for the six blood-based biomarkers of dementia as well as melatonin and cortisol.

Rhythmicity of melatonin and cortisol did not differ between groups. P-Tau217 and GFAP (p<0.05) and BD-Tau (p<0.10) were elevated in PLWA. Significant time-of-day variation was observed for BD-Tau as well as the other five biomarkers of dementia, with peak levels coinciding with the sleep period. Group by time-of-day interactions were observed for p-Tau217 and Aβ42. Average BD-Tau, p-Tau217, and GFAP concentration were negatively correlated with memory function when averaged over the 15 assessments, but these correlations became less robust when the number of repeat assessments was smaller. Mood and daytime sleepiness did not correlate with any of the biomarkers. The relationship between p-Tau217 concentration and memory function was strongest for blood samples collected during the sleep period.

These data have implications for our understanding of the association between blood-based biomarkers and daytime function and the design of research protocols. The observation that BD-Tau and other plasma biomarkers vary with time of day with highest levels during the sleep period is consistent with sleep or circadian rhythmicity mediated facilitation of overnight clearance of these proteins from brain to blood.

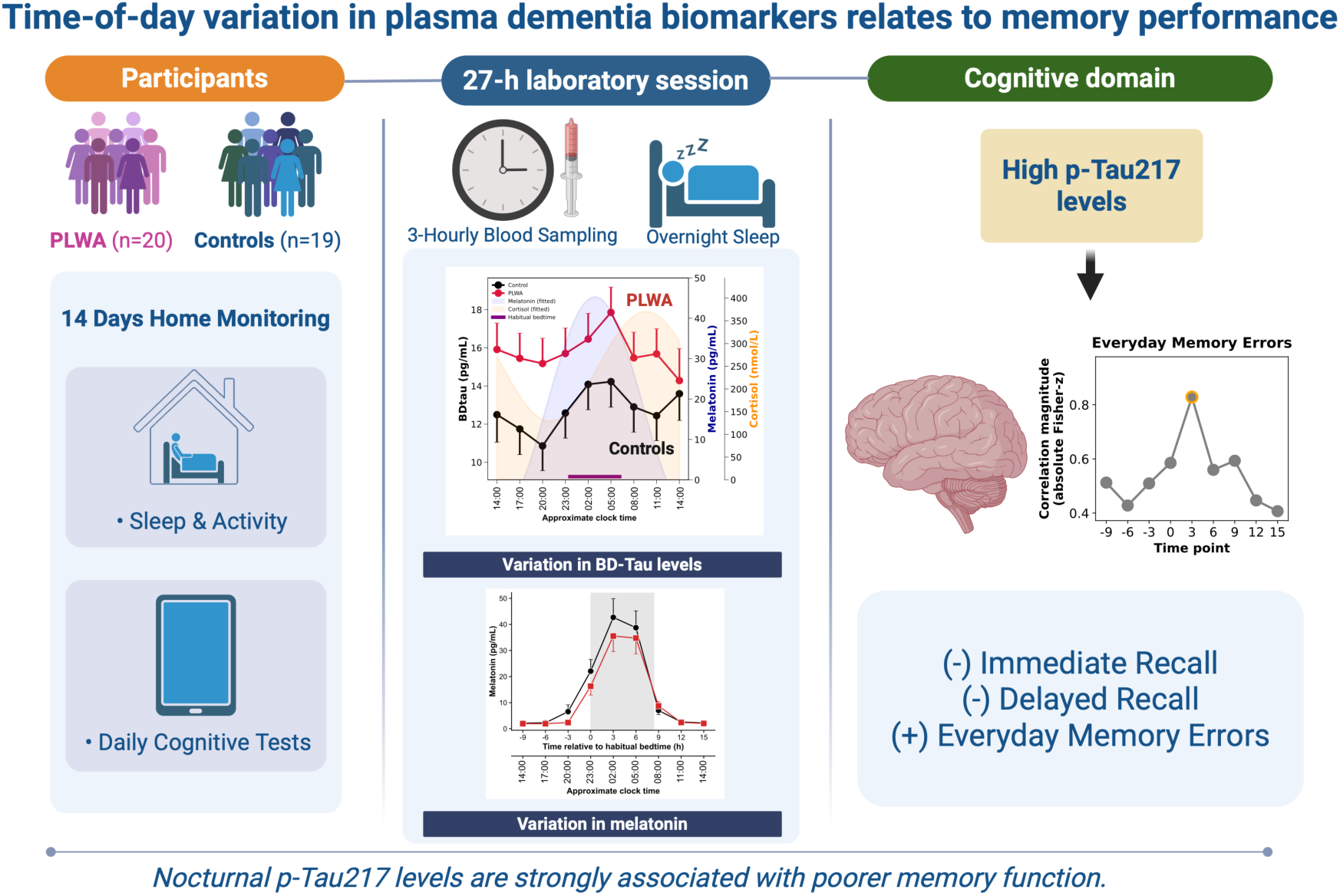
Graphical Abstract.

## Introduction

Alzheimer’s disease (AD) is characterised by neuropathological changes, including amyloid-β (Aβ) aggregation and tau hyperphosphorylation, which begin well before clinically significant alterations in cognitive function are observed^1–3^. As such, blood-based biomarkers that reflect these pathologies including brain derived tau (BD-Tau), phosphorylated tau (e.g., p-Tau217), Aβ isoforms, neurofilament light chain (NfL), and glial fibrillary acidic protein (GFAP), have emerged as tools for minimally invasive early diagnosis, population screening, monitoring disease progression, and evaluating therapeutic efficacy.

Many blood-based biomarkers of dementia, however, originate from both brain and peripheral tissues which may compromise their specificity for brain pathology. BD-Tau comprises isoforms of tau that are brain-specific and show specificity to AD-type neurodegeneration^4^. Indeed, elevated levels of BD-Tau can be used to identify Aβ-positive individuals at risk of rapid cognitive decline and atrophy^5^.

The validity and usefulness of blood-based biomarkers of dementia have most often been assessed by comparison to biomarkers from cerebrospinal fluid (CSF). Biomarker levels in CSF and plasma are strongly correlated although plasma levels are an order of magnitude lower than those in CSF (e.g.,^6^). The validity and usefulness of blood-based biomarkers of dementia may also be assessed by investigating their association with brain function. For example, p-Tau217 was shown to be able to predict memory deficits in those living with AD outperforming p-Tau181 and p-Tau231^7^. Increases in p-Tau217 levels over time have been shown to associate with cognitive decline in memory, language, attention, and executive function, as well as disease progression^8^. Furthermore, p-Tau217 levels have been shown to predict levels of agitation and aggression in people living with AD (PLWA)^9^.

Several covariates that affect biomarker concentration in plasma and thereby reduce their predictive or diagnostic value, have been identified. These include body mass index (BMI), blood volume, kidney function, or comorbid conditions^10–12^. Intra-individual variability in biomarker levels, may also be driven by factors such as activity levels, posture, sleep patterns, meal consumption or time of day/circadian rhythmicity^13^. Disruption in overt diurnal rhythmicity and sleep are a hallmark of AD progression, and bidirectional relationships between AD and disturbance in sleep and circadian rhythmicity, which may be indexed by rhythmicity in endocrine markers such as melatonin and cortisol^14,15^, have been frequently posited^16^.

There is emerging evidence that blood-based biomarkers of dementia fluctuate across the 24-h period^17^. However, most of these studies only compared an evening vs a morning samples rather than characterising a full 24-hour profile (e.g.,^6,18^). One study took two-hourly plasma and CSF samples in five cognitively normal participants for 36 hours under conditions of sleep and sleep deprivation^19^. Under conditions of sleep deprivation, levels of Aβ40, Aβ42, p-tau181 and unphosphorylated tau-217 increased in CSF and decreased in plasma suggesting a reduction in brain clearance mechanisms in the absence of nocturnal sleep^19^.

Recently, we investigated time-of-day variation in blood-based biomarkers of dementia by taking time series blood samples at 3-hourly intervals across the 24-hours day under controlled conditions in our clinical research facility. In a small population of older adults of both cognitively intact individuals and those with mild AD, we demonstrated, that there was significant diurnal variation in plasma levels of p-Tau217, Aβ40, Aβ42, and NfL, with lowest values in the early morning and peak values in the early evening/overnight^20^.

Quantifying the extent of and identifying the factors that drive time of day variation in these biomarkers and how well they correlate with cognitive status, will be crucial in establishing protocols for their clinical implementation. Of all the blood-based biomarkers of dementia, to date, p-Tau217 has been heralded as the most accurate tool for predicting cognitive decline^21^ as well as monitoring treatment efficacy^22,23^. One unresolved question is whether p-Tau217 levels and other biomarkers, such as BD-Tau, reflect more general aspects of brain function (e.g., alertness, mood, speed) or are only associated with more specific aspects of brain function such as short-term memory. In most studies to date, associations between biomarkers and brain function were based on single cognitive assessments^7,24^. It therefore also remains to be established to what extent repeat assessments of brain function are required to obtain a reliable estimate of these associations.

The primary aims of this study were to: 1) further quantify the diurnal variation in BD-Tau and other blood-based biomarkers; 2) investigate the association between biomarkers and a range of daytime function measures and sleep; 3) investigate whether clinically meaningful biomarker-brain function relationships can be captured from a single blood sample and whether it matters at which clock time the sample is taken, 4) investigate whether one assessment of brain function is sufficient to establish associations between biomarkers and brain function or whether repeated assessments are required. The results inform the development of optimal sampling protocols for both research and clinical settings and aid the interpretation of the results.

## Materials and Methods

A subset of the blood-based biomarker data has been previously published^20^. Here we extend the sample size; include BD-Tau, melatonin, and cortisol data; and combine the biomarkers with measures of waking function and sleep, none of which have previously been reported.

### Participants

Participants were people living with mild or prodromal Alzheimer’s disease (PLWA), their supporting study partner (family member, caregiver or friend), and cognitively intact older adults. The eligibility criteria have been described in detail elsewhere^20,25,26^. Diagnosis of AD was based on clinical history, cognitive tests, and computerised tomography (CT) or magnetic resonance imaging (MRI) scans. We enrolled 20 PLWA (7F) aged 72.2 ± 6.0 years (mean ± SD) who had a Standardised Mini Mental State Examination (sMMSE) score ≥ 23 and lived in the community. We also enrolled 19 control participants (10 F; 68.9 ± 9.8 years) who were either study partners (≥18 years) or cognitively intact older adults (50-85 years) and had an sMMSE score ≥ 27. The participants were recruited by the Surrey Clinical Research Facility (cognitively intact) or Surrey and Borders Partnership NHS Foundation Trust (SABP) memory services (PLWA and study partners).

### Ethics and governance

The study received a favourable opinion from an NHS ethics committee (22/LO/0694) and was registered as a clinical study on the ISRCTN (International Standard Randomised Controlled Trial Number) registry (ISRCTN10509121). The study was conducted in accordance with the Declaration of Helsinki and guided by the principles of Good Clinical Practice. All personal data were handled in accordance with the general data protection regulations (GDPR) and the UK Data Protection Act 2018. Participants provided written informed consent prior to any study procedures being performed and were compensated for their time and inconvenience.

### Study Design

The protocol has been described previously^20,25^. Following a screening visit to assess eligibility, participants completed a 14-day at-home data collection period during which they completed a daily sleep diary^27^ and had their sleep monitored with wearable and contactless technology including the Withings Sleep Analyser (WSA; Withings, France)^28^. Participants were instructed that each morning, ∼2 hours after waking, they were to complete an electronic waking function assessment on the Cognitron platform^29^. The measures of daytime function assessed were: Karolinska Sleepiness Scale^30^ (KSS), Bond and Lader scales^31^, simple reaction time^29^, choice reaction time^29^, immediate memory word recall^32^, delayed memory word recall^32^, and self-reported everyday memory errors^32,33^.

At the end of the two-weeks, participants attended the UK-DRI Clinical Research Facility at the University of Surrey for a 27-h residential session with an extended 10-h period in bed. During this session, blood samples were collected at 3-hourly intervals for 24 hours beginning 9 h before habitual bedtime and ending 15 h after habitual bedtime. Light levels were controlled and set at ∼235 lux in the vertical plane at 1.6 m. Mealtimes were standardised as follows: lunch was ∼9.5 h before habitual bedtime, dinner was ∼4 h before and breakfast was ∼1.5 h after habitual waketime. Blood samples were collected via a cannula into Lithium Heparin and K2 EDTA vacuettes, centrifuged within 10 min of collection at 4 °C at 1620 × g for 10 min, and the plasma fraction separated and stored at −80 °C. The samples were analysed for melatonin and cortisol using liquid chromatography mass spectrometry (LCMS) (Chrono@work, The Netherlands). The blood-based biomarkers of dementia were analysed as follows: a) Neuro 4-PlexE assay kit (Quanterix, Billerica MA) for Aβ40, Aβ42, GFAP, and NfL; b) ALZpath Simoa p-Tau217 and BD-Tau.

### Data Analysis

Sleep variables from the WSA were extracted from the automated analysis and averaged across the 15 days of the study (14 days at home and the laboratory session). Variables d included in the analyses were: total sleep time (TST), wake after sleep onset (WASO), sleep efficiency (SE) and apnoea-hypopnea index (AHI). Validity of these measures has previously been reported in middle-aged adults^34^, older adults^35^, and those with sleep apnoea^36^.

For the measures of daytime function, a single value was extracted for each measure for each day and log transformed as necessary: a) KSS – raw score, b) Choice reaction time and Simple Reaction time – reaction time in msec (mean and standard deviation), c) Everyday memory errors – number of errors, d) Immediate recall and Delayed recall – number of words correctly recalled. We also extracted measures of Alertness, Contentedness, and Calmness scores from the Bond & Lader visual analogue mood scales and these were transformed into z-scores.

The timing of melatonin production was quantified as the midpoint between the upward and downward limb crossings of the mean value (e.g.,^37^). The individual timings of sample collection were then calculated relative to the melatonin midpoint for each participant and grouped into bins. Nine bins were created where the centre of the bin was based on the distribution of melatonin phases per sample number to ensure that almost all samples of a particular sample number fell within a bin. In addition, the mean level was calculated for each biomarker profile.

### Statistical Analysis

For the purposes of the current analysis, participants were divided into two groups: 1) PLWA, and 2) Control, which comprised the study partners and cognitively intact older adults’ data sets.

To assess variation of measures with time, linear mixed models were run using PROC MIXED (SAS v9.4, SAS Institute Inc) in which participant was the random effect with factors Time and Group. For the biomarkers, the factor Time was time-of-day (melatonin bin) and for the measures of daytime function the factor Time related to the day number in the study. To investigate whether associations between measures of daytime function and blood-based biomarkers depend on time of day at which the biomarkers samples were collected we conducted an additional mixed model analysis. In this model the biomarker Timepoints (categorical variable) and Daytime measures (using the average over 15 days) were independent variables, and the dependent variables were the Biomarker levels with Participant as the random effect. A significant interaction between Timepoints and Daytime measures indicates that the association between the biomarker and the daytime function measure depends on the time of sampling. All the simple (only Time and Group as factors) linear mixed models were repeated with a more complex structure including the covariates age, sex, BMI, AHI, and PSQI.

To determine the intra– vs inter-individual variation for both biomarker levels and daytime function, intraclass correlation coefficient (ICC_1,1_) analysis was performed in R (v. 4.2.2, R Core Team 2022, Psych library, ICC function with two-way random effect) using the dependent variable (biomarker levels or cognitive measure) as “response” and participant ID as “subject”. ICC values were interpreted as: below 0.50 indicate poor reliability, values between 0.50 and 0.75 indicate moderate reliability, values between 0.75 and 0.90 indicate good reliability, and values above 0.90 indicate excellent reliability^38^.

To explore the relationship between daytime function and biomarker levels, Pearson correlations were performed between individual mean values for daytime function (mean across 15 consecutive days) and the biomarkers (mean across nine timepoints taken over 24 h). Subsequently, correlation magnitudes were converted into absolute Fisher’s z-scores. To investigate how associations between biomarkers and daytime function measured depended on the number of blood samples and the number of function measures we proceeded as follows. Correlations were calculated between each of the biomarker timepoints (as well as the mean of the 9 timepoints) with each daytime function measure. Estimates of daytime function measures were based on backward cumulative averaging i.e., for day 1 (laboratory session) this was the day 1 value, for day –1 it was the mean of day –1 and 1, for day –2 it was the mean of days –2, –1, and 1 etc until the mean of all 15 days was calculated. Pearson correlation p-values were corrected for multiple testing using the Benjamini–Hochberg false discovery rate (FDR) procedure^39^.

Finally, to assess the reliability of the daytime function variables, for each one a single measurement ICC was estimated and then the reliability of an averaged composite was computed using the Spearman-Brown formula^40^ given below.

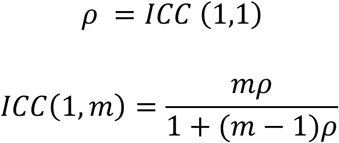

(m is the number of parallel repeats averaged to form a composite score; p is the single reliability measure).

This approach assumes that each repeat has similar variance. From this analysis, we identified the smallest number of repeats needed for an ICC to achieve ≥ 0.90.

## Results

### Participants

20 PLWA and 19 Control participants (n = 12 study partners) completed the study. A summary of the participant demographics is provided in Supplemental Table 1. The two groups were similar in age (PLWA: 72.2 ± 6.0; Controls: 68.9 ± 9.8 years) and BMI (PLWA: 25.6 ± 6.6; Controls: 26.7 ± 4.0). The PLWA had significantly lower s-MMSE scores (PLWA: 27.2 ± 1.6; Controls: 28.9 ± 0.9), lower activities of daily living score (PLWA: 7.3 ± 1.3; Controls: 8.0 ± 0.2), increased depression (PLWA: 4.0 ± 2.5; Controls: 1.7 ± 1.4), and less morning type tendencies (PLWA: 58.1 ± 6.8; Controls: 62.3 ± 11.4).

### Time of day variation in biomarkers

The melatonin and cortisol profiles showed the expected time-of-day variation with peak levels overnight and in the morning, respectively (Figures 1 and 2). The hormone profiles did not significantly differ between groups.

**Figure 1.**
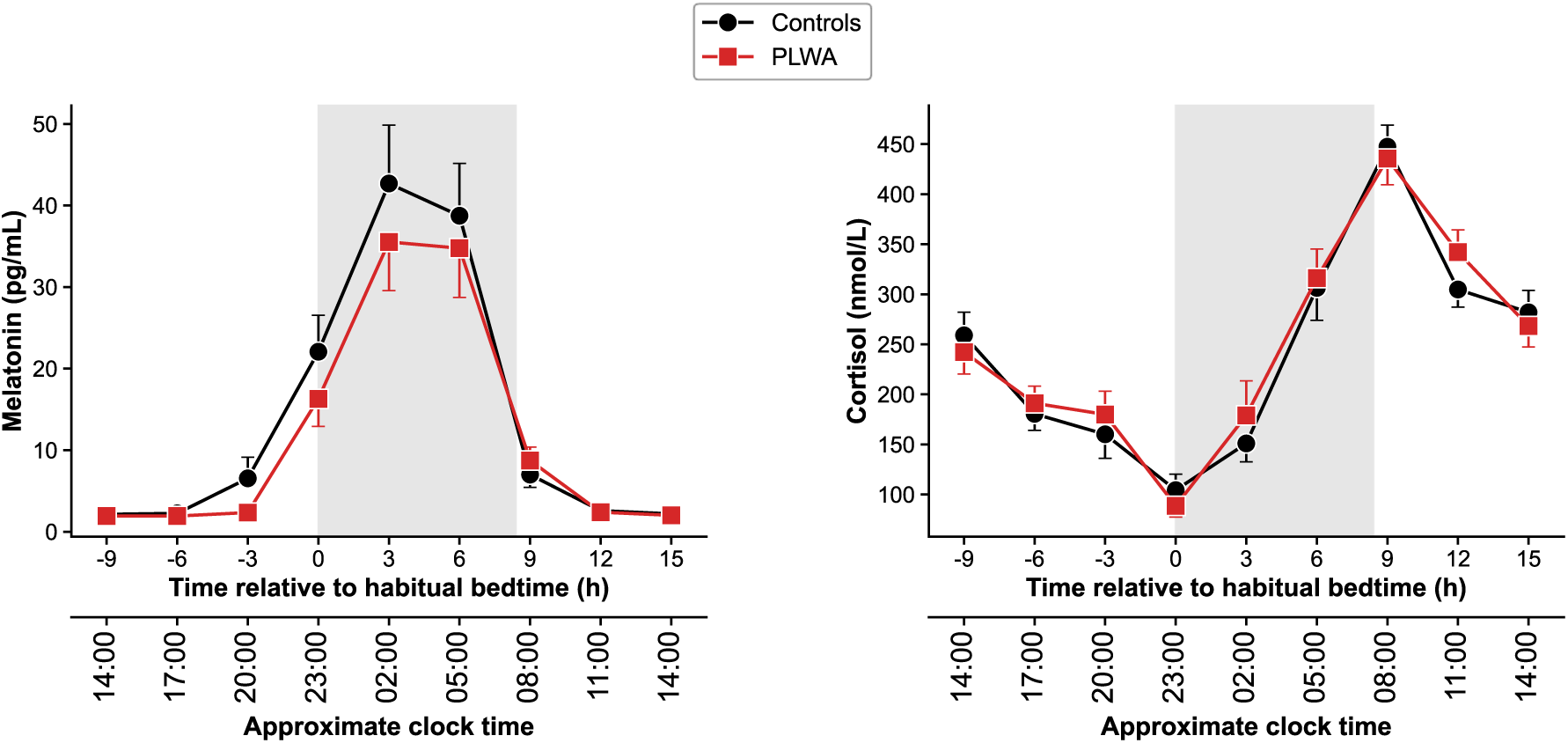
Melatonin (left panel) and cortisol (right panel) profiles (mean ± SE) over a 24-hour period in PLWA (red lines) and controls (black lines). Grey area indicates the average timing of the sleep periods.

**Figure 2.**
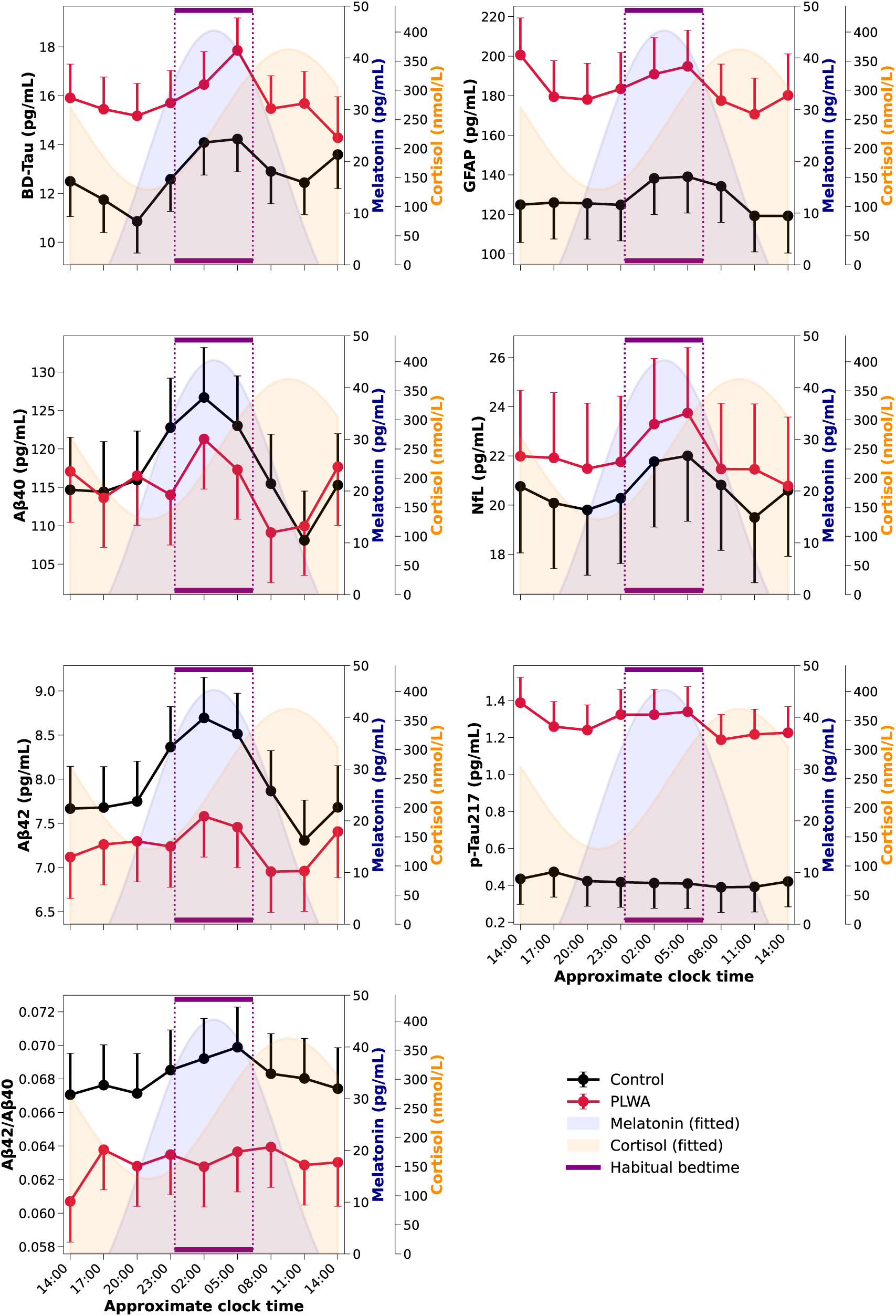
LS-Means and SE for the 24-hour profiles of blood-based biomarkers of dementia in PLWA (red lines) and controls (black lines) superimposed on average melatonin (blue shading) and cortisol (yellow shading) profiles. Average habitual sleep period is indicated by the purple horizontal lines.

Significant group differences were observed for p-Tau217 (p < 0.001) and GFAP (p < 0.05) with higher levels of both markers in PLWA. For BD-Tau, there was a trend for higher levels in PLWA (p = 0.085). BD-Tau exhibited significant time of day variation as did the remaining five biomarkers as well as the Aβ42/40 ratio. For all biomarkers highest levels coincided with the sleep period (Figure 2, Supplemental Table 2, and Supplemental Figure 1). Significant interactions between time-of-day and group effects were observed for p-Tau217 (p < 0.01) and Aβ42 (p < 0.05) (Figure 2 and Supplemental Table 2). When the more complex model was run, the group and time effects remained, and significant effects were observed for age (BD-Tau, Aβ40, Aβ42, GFAP, NfL, cortisol), AHI (BD-Tau, Aβ40), BMI (Aβ40, Aβ42, Aβ42/Aβ40), sex (cortisol), and PSQI (cortisol). Correlation analyses to assess how the average concentrations of each biomarker correlated with other biomarkers revealed 15 significant associations of which 13 were positive. The two negative correlations were for Aβ42/Aβ40 with p-Tau217 and GFAP (Supplemental Table 3).

ICC analyses (Supplemental Table 4) revealed that the within-individual (diurnal) variation compared to the between individual variation for the dementia plasma biomarkers was rather small (i.e. large ICC values, total sample: 0.782 – 0.973) which translates to good-excellent reliability of a single sample. By contrast, for plasma melatonin and cortisol the within-individual (diurnal) variation was larger compared to the between individual variation (small ICC values; for total sample: 0.121-0.128; poor reliability), and this for both the complete dataset and the controls and PLWA separately.

### Measures of memory, mood, and daytime sleepiness differs between PLWA and Control

Measures of daytime function across the study are shown for the two groups in Figure 3. Significant between groups differences were observed with PLWA having more memory errors, reduced performance in both immediate and delayed recall (p < 0.001), being sleepier and less content, and having more variable choice reaction time responses (p < 0.05) (Supplemental Table 5). The group effects persisted for the memory tasks but not for contentedness, choice reaction time, or sleepiness when the more complex model with additional covariates was run. Significant effects of time (days) were observed for a number of variables which may, in part, reflect learning effects: a) Choice Reaction Time and Simple Reaction Time: mean reaction time and standard deviation, b) Everyday memory errors: total errors c) Delayed Recall: number correct, d) Calmness (Supplemental Table 5).

**Figure 3.**
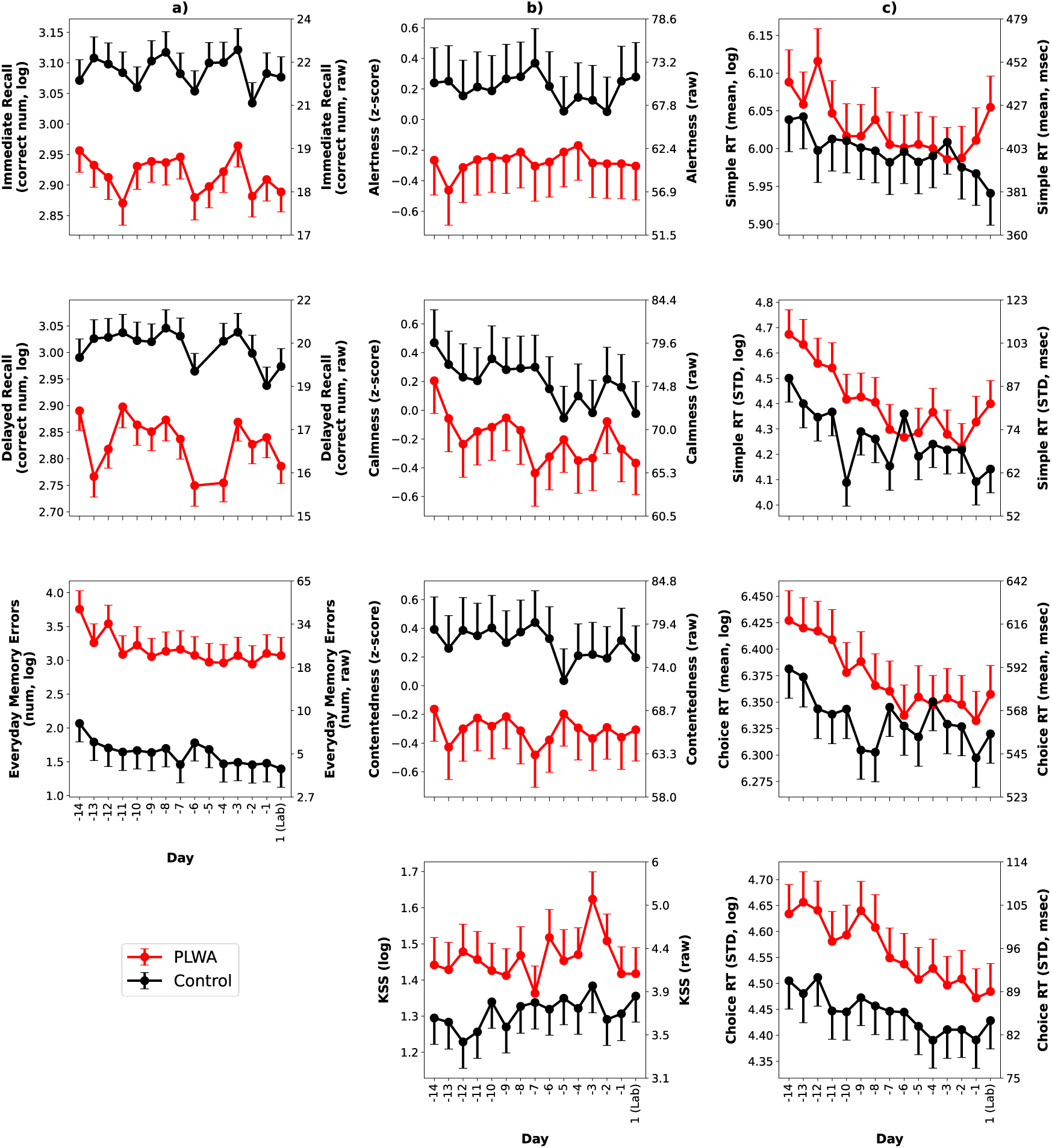
LS-Means and SE for the measures of daytime function in PLWA (red lines) and controls (black lines) over a two-week period (–14 to –1) at home and 1 day in the clinical research lab.

### Relationship between biomarkers and measures of daytime function are domain specific

Figure 4 (upper panel) shows that the strength of the relationship depended upon the biomarker and the cognitive domain, with the strongest associations observed for measures of memory and the lowest for daytime sleepiness. Significant (surviving FDR) negative associations were observed between: BD-Tau and delayed recall (p<0.01); p-Tau217 and both immediate and delayed recall (p<0.001); GFAP and both immediate and delayed recall (p<0.01) (Figure 4 lower panel, and Supplemental Table 6).

**Figure 4.**
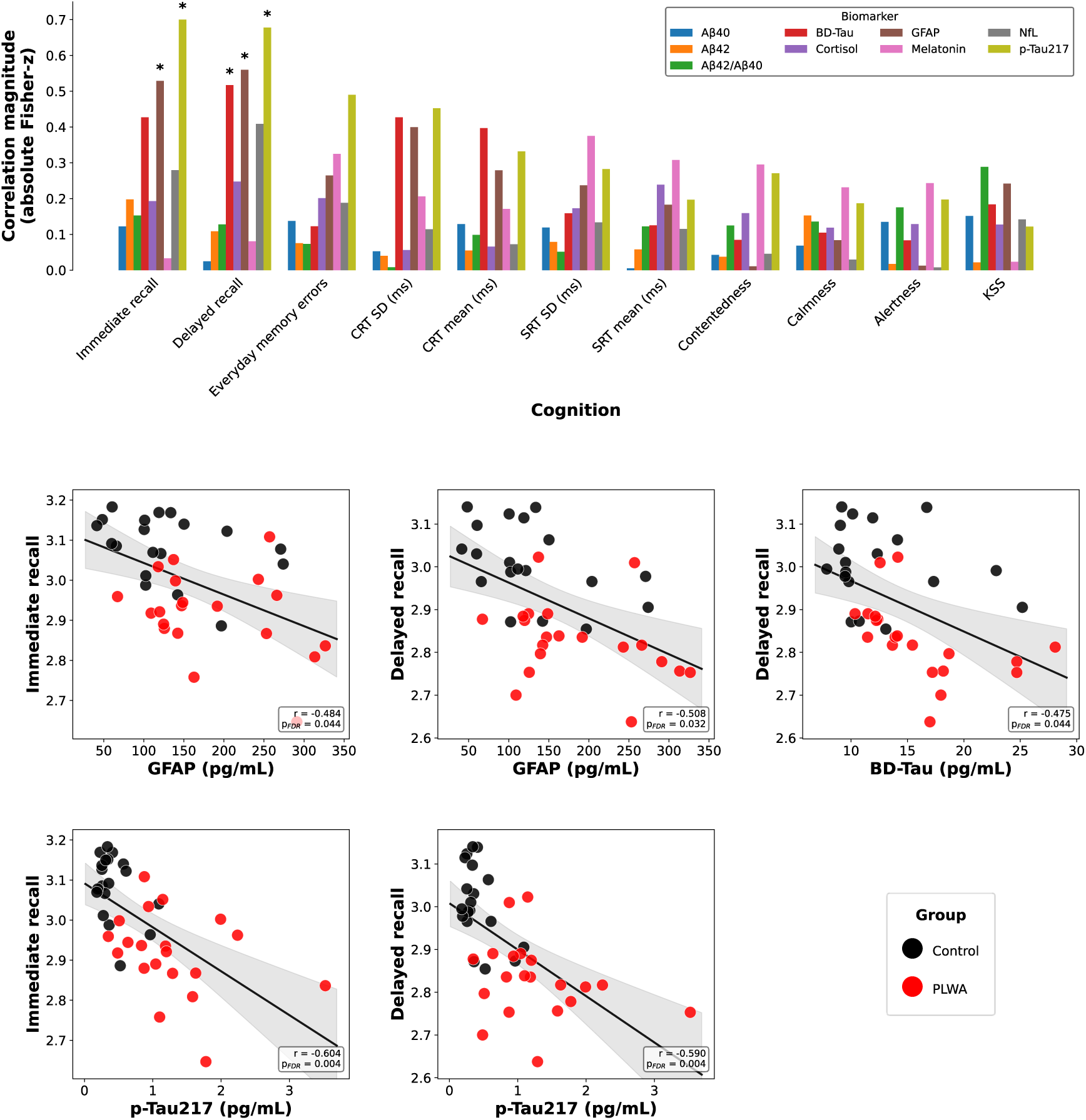
Correlations between biomarker levels (mean of nine samples) and measures of daytime function and sleep (mean of 15 days). Upper panel: correlation magnitudes for eight blood-based biomarkers and measures of daytime function. Significant associations are indicated with an asterisk. Lower panel: significant individual correlations for p-Tau217, GFAP and BD-Tau.

### Relationship between biomarkers and measures of sleep

None of the associations between biomarkers and measures of sleep remained significant after FDR correction (Supplemental Table 7). For all participants combined, the following nominally significant associations were observed. Aβ40 correlated positively with deep sleep duration (p < 0.05) and time in bed (p < 0.05); Aβ42 correlated positively with deep sleep duration (p < 0.05 and NfL correlated positively with time in bed (p < 0.05). Average melatonin concentration correlated positively with REM duration (p < 0.01) and negatively with light sleep duration (p < 0.05); When the correlation magnitudes were reviewed (Supplemental Figure 2), highest associations were observed for time in bed with NfL and Aβ40, and for AHI and melatonin, whereas lowest associations were observed for the biomarkers with sleep efficiency and duration of wake.

### Associations between biomarkers and daytime function depend on the timing of blood sampling

Mixed model analysis (Supplemental Table 8) revealed significant associations between biomarker levels and cognitive measures (averaged over 15 assessments) for: BD-Tau and CRT_STD (p<0.01); CRT_mean (p<0.05), delayed recall (p<0.01), immediate recall (p<0.05); p-Tau217 and immediate recall, delayed recall, everyday memory errors, and CRT_STD (p<0.01), and CRT_mean (p<0.05); GFAP and delayed recall, immediate recall (p < 0.01), and CRT_STD (p<0.05);) NfL and delayed recall (p<0.05). Significant interactions between time and cognitive measure indicate that the strength of the associations between cognitive measures and blood-based biomarkers were dependent upon the time of day at which the blood sample was taken. Significant interactions were observed for: BD-Tau and CRT-mean (p<0.001); p-Tau217 and delayed recall, immediate recall, and everyday memory errors (p<0.01); GFAP and SRT_STD (p<0.01); NfL and alertness (p<0.05), and AB42/AB40 and delayed recall and immediate recall (p<0.05).

For p-Tau217 the strongest associations with memory measures were observed for samples collected during the sleep period. (Figure 5a), and significant associations were observed with only a single assessment of daytime function (Supplemental Figure 3). When we analysed how the strength of these correlations varied with number of assessments over which memory function was averaged, it emerged that more assessments in general resulted in a more significant association (Figure 5b).

**Figure 5.**
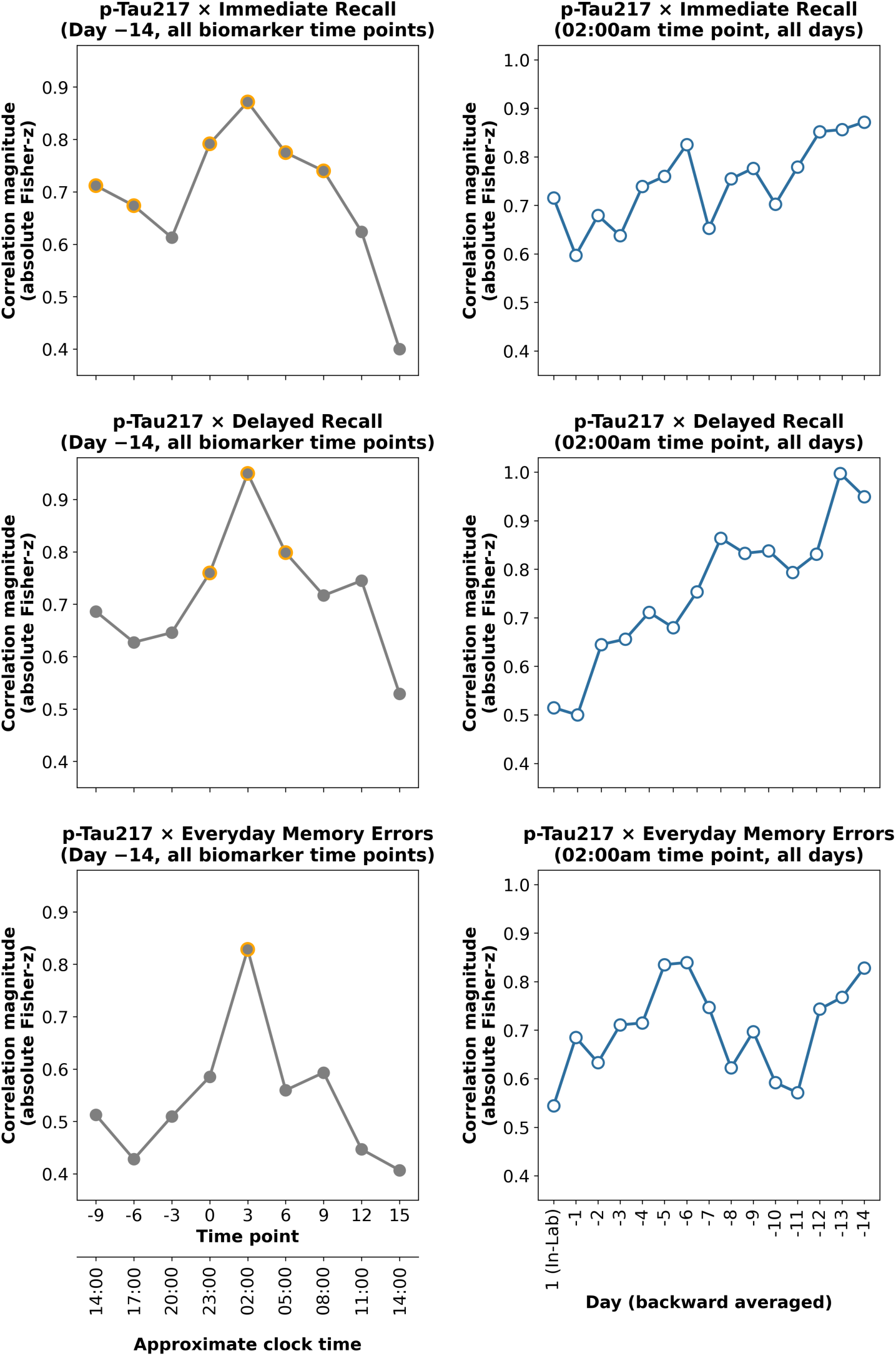
Left panel: Strength of the association (Pearson r coefficient values transformed into the absolute values of Fisher-z scores) between p-Tau217 levels and memory varied across the 24 h day. Orange circles indicate the most significant associations for that measure of daytime function: a) & b) p < 0.001, c) p<0.01. Right panel: Correlation magnitudes between the p-Tau217 levels at 2am and the cognitive measure backward averaged across study days.

### Reliability of assessments

To quantify the reliability of each of the daytime function measures we conducted Intraclass correlation analyses. ICC analyses for the daytime function measures (Supplemental Table 9) revealed that for seven of the 11 measures, the ICC values were lower in PLWA implying greater within-individual (day-to-day) variation in this group. There was high within-individual (day-to-day) variation compared to between-individual variation for measures of memory recall in both PLWA (ICC: 0.289-0.391, and controls (ICC: 0.358-0.388, poor reliability). For subjective measures of mood, the within-individual variation was relatively small for control participants (0.524-0.801(moderate to good reliability)) but slightly larger for PLWA (ICC: 0.408-0.809; poor to good reliability). For reaction time measures, the range of ICC values was larger in PLWA (ICC: 0.298-0.746) than in Controls (ICC: 0.488-0.779).

We then conducted analyses to determine the number of assessments required to obtain a reliable assessment of this measure for the whole population and separately for PLWA and Controls (Supplemental Figure 4). For all participants combined, the most reliable measure requiring the least number of days was the subjective assessment of memory (Everyday Memory Errors) which required two days. The least reliable was SRT_STD which is a measure of consistency of reaction time within the test, which required 11 days. Subjective sleepiness (KSS), Delayed recall and Immediate recall required 10, nine and eight days, respectively to stabilise. When reliability was assessed separately for PLWA and Controls, more days were required for PLWA for Delayed Recall (23 vs 15 days), SRT_STD)14 vs 10 days), CRT_STD (22 vs 6 days), and KSS (14 vs 9 days). For Immediate recall, PLWA required 15 days compared to 17 days for Controls.

## Discussion

The results establish that BD-Tau and five other blood-based biomarkers of dementia exhibit time-of-day variation, with highest values during the sleep period. The day-to-day variation was relatively small compared to the inter-individual variation.

Levels of BD-Tau, p-Tau217, and GFAP associated with measures of memory function but not with other measures of daytime function. The associations between the levels of biomarkers and memory were strongest for blood samples collected during the nocturnal sleep episode and were strongest when all 15 assessments were used to quantify memory function. The findings have implications for our understanding of the association between blood-based biomarkers and daytime function and the design of research protocols.

The mechanisms underlying the time-of-day variation in plasma biomarker levels remains unclear. Some previous studies exploring time-of-day variation in plasma or CSF biomarkers did not observe differences across the 24-hour day for the markers we used (e.g., p-Tau217, GFAP, NfL)^6^. However, rather than using timeseries sampling these studies simply compared levels in a morning vs a single evening sample where according to our data, and depending on the exact timing of the sample, levels may indeed not be significantly different. The current data establish that difference between morning and evening samples are in general much smaller than differences between middle of the ‘night’ and morning and evening samples.

Time of day variation in biomarker levels in plasma may reflect diurnal variation in production or in clearance from the brain to and from blood. Clearance mechanisms implicated are the glymphatic system, transport via the blood-brain barrier or reflect diurnal ‘clearance’ from blood, i.e. peripheral degradation. Although the profile of p-Tau 217 was slightly different from the other profiles, a general descriptor of the diurnal variation observed in the present study is that the time course of diurnal variation was similar for all biomarkers including BD-tau, which is produced exclusively in the brain. This implies that peripheral production is an unlikely driver of the diurnal variation and that this variation is driven by one global mechanism. Brain clearance mechanisms may be modulated by sleep or circadian mechanisms. Glymphatic influx, clearance and CSF drainage has been proposed to be under circadian control in mice with peak activity during the rest period^41^ implicating the circadian system in driving time of day variation in plasma biomarker levels. In support of sleep-dependent mechanisms, it has recently been demonstrated in mice that endocytosis in brain endothelial cells of the blood-brain-barrier is enhanced during sleep which enables the removal of brain-derived waste into circulation. Blocking these endocytosis processes results in increased sleep suggesting that if brain clearance is insufficient then the brain compensates through increased sleep^42^ (Li et al., 2026). A three-way, randomised crossover study in healthy young adults (20-40 years) explored the impact of sleep vs a total night of sleep deprivation on morning and afternoon CSF levels of p-Tau181, GFAP, NfL, orexin, Aβ38, Aβ40 and Aβ42^43^. Morning CSF levels of Aβ and p-tau181 were consistently lower after sleep compared to after a night of sleep deprivation or the afternoon suggesting that a sleep-driven mechanism reduced these CSF levels perhaps due to enhanced solute motility^43^. Recently, a randomised crossover design in older adults compared biomarker levels following a night of sleep or night of sleep deprivation^44^. Plasma biomarker levels were higher following the night of sleep, and this was proposed to result from increased glymphatic clearance from CSF to plasma mediated by a sleep-active clearance processes facilitated by lowered brain parenchymal resistance. In a study, in which the response to presence or absence of sleep was assessed in both CSF and plasma with a high temporal resolution sampling protocol, demonstrated that in the absence of sleep levels of Aβ40, Aβ42, p-Tau181 and unphosphorylated Tau-217 increased in CSF and decreased in plasma. The data were interpreted as in support of sleep facilitated clearance from brain to blood^19^ (Liu et al., 2023). We concur that this is for now the most parsimonious explanation for our observations.

The magnitude of the diurnal variation was modest compared to the inter-individual variation which suggest that it may not significantly impact the usefulness of the biomarkers for diagnostic purposes although the diurnal variation may be relevant in situations in which the detection of small changes in biomarker levels within an individual are required. The observation that levels of BD-Tau, p-Tau217, and GFAP are related to memory function, but not to other aspects of daytime function such as mood or sleepiness, speaks to the specificity of these biomarkers for impairment of brain function central to Alzheimer’s disease. Associations between p-Tau217 and memory function have been previously reported. In a cross-sectional study of more than 700 adults (cognitively impaired and unimpaired), p-Tau217 was found to outperform other p-Tau markers (181 and 231) in terms of identifying memory impairment and, in one cohort, impairment of executive function^7^. A recent study assessing cognitively unimpaired and those with mild cognitive impairment demonstrated that p-Tau217 levels alone were associated with a greater decline in episodic/semantic memory and processing speed^45^. Interestingly, when the p-Tau217 results were combined with GFAP levels, the observed decline in cognitive performance extended to most cognitive domains including global cognition and executive function suggesting the utility of a multi-biomarker panel to characterise cognitive status. When we conducted a similar analysis and combined p-Tau217 and GFAP we found that CRT variability and everyday memory errors were the measures that were most sensitive to combined biomarkers.

Surprisingly, the associations between p-Tau217 and other biomarkers were strongest for blood samples taken overnight, but a simple explanation for this observation is not available. The analyses of the inter-individual variation in daytime function assessments and the observation that this variation is greatest for objective memory assessments and larger in PLWA is in accordance with a previous study^32^. Variation of reliability between the various assessments of brain function, and the observation that this reliability may be lower in PLWA has implication for the design of research protocols to investigate association between biomarkers and brain function. Repeated assessments of brain function are more likely to reveal association between blood-based markers and brain function than single assessments.

## Limitations

The main limitation of this work is the small sample size, but this is compensated by the intense timeseries approach with multiple blood samples collected over a 24-hour period (biomarkers) and daytime function assessments over multiple days.

## Conclusion

Blood-based biomarkers for dementia exhibit time-of-day variation with highest levels during the nocturnal sleep period and lowest levels in the morning. The relationship between levels of the biomarkers and cognitive function is domain specific, predominantly relating to memory, depends on the timing of the sample and the reliability of the cognitive assessment.

## Acknowledgements

We thank Surrey Clinical Research Facility for support with recruitment and residential sessions, in particular Kat Pizzoferro, for her oversight of sample collection and processing. We also thank Jasmine Ramble for assistance with data curation. We thank Surrey & Borders Partnership Trust for their role in recruitment and screening of participants. We also thank all the Cognitron Team for their support on the digital platform. Thanks to Dr Ullrich Bartsch for discussions and comments on the manuscript.

## Funding

This work is supported by the UK Dementia Research Institute Core Award (CF2023\7 and UKDRI-7206 (DJD) through UK DRI Ltd, principally funded by the UK Medical Research Council, and the NIHR Oxford Health Biomedical Research Centre [NIHR 203316], and the UK Dementia Research Institute at UCL (UKDRI-1003).

## Competing Interests

DJD received equipment from Somnofy and is a consultant to Boehringer-Ingelheim, Danisco Sweeteners OY, and AstronauTx. The remaining authors declared that this work was conducted in the absence of any commercial or financial relationships that could be construed as a potential conflict of interest.

## Data Availability

The raw data supporting the conclusions of this article will be made available by the authors, without undue reservation.

## Notes

### Author Declarations

The study received a favourable opinion from an NHS ethics committee, London – City and East Research Ethics Committee (REC) (22/LO/0694)

