## Supplemental Tables and Figures for "Diurnal variation in brain-derived tau and five other blood-based biomarkers for dementia and their association with cognitive performance"

### Supplementary Material

**Supplemental Table 1.** Participant Demographics.

| Participant Group | Total Sample | PLWA | Control |
|---|---|---|---|
| N | 39 | 20 | 19 |
| **Sex - Males (%)** | 22 (56.41) | 13 (65.00) | 9 (47.37) |
| **Sex - Females (%)** | 17 (43.59) | 7 (35.00) | 10 (52.63) |
| **Age (+/- SD)** | 70.67 (8.10) | 72.2 (6.04) | 68.89 (9.76) |
| **BMI (+/- SD)** | 26.38 (5.61) | 25.56 (6.63) | 26.69 (4.09) |
| **Race - White (%)** | 33 (84.62) | 17 (85.00) | 16 (84.21) |
| **Race - Asian (%)** | 4 (10.36) | 2 (10.00) | 2 (10.53) |
| **Race - Indian mixed** | 1 (2.56) | 1 (5.00) | 0 (0.00) |
| **Race - Asian/South American (%)** | 1 (2.56) | 0 (0.00) | 1 (5.26) |
| **Education level - None (%)** | 3 (7.69) | 4 (20.00) | 1 (5.26) |
| **Education level - GCSE/ O-level (%)** | 6 (15.38) | 7 (35.00) | 3 (15.79) |
| **Education level - AS/A levels / L3 diploma** | 7 (17.95) | 3 (15.00) | 4 (21.05) |
| **Education level - Bachelor's Degree or Equivalent (%)** | 15 (38.46) | 5 (25.00) | 9 (47.37) |
| **Education level – Master's Degree or Equivalent (%)** | 1 (2.56) | 0 (0.00) | 1 (5.26) |
| **Education level – Other (%)** | 7 (17.95) | 1 (5.00) | 1 (5.26) |
| **sMMSE (+/- SD)** | 27.95 (1.62) | 27.16 (1.61) | 28.89 (0.94)* |
| **PSQI (+/- SD)** | 4.38 (3.44) | 4.58 (3.83) | 4.37 (3.09) |
| **AHI (+/- SD)** | 16.81 (13.10) | 20.17 (14.74) | 13.27 (10.36) |
| **PLMI (+/- SD)** | 12.30 (20.78) | 15.73 (26.10) | 8.69 (12.85) |
| **H-O (+/- SD)** | 60.19 (9.51) | 58.11 (6.83) | 62.26 (11.40) |
| **NART (+/- SD)** | 16.59 (9.95) | 18.85 (10.39) | 14.21 (9.13) |
| **ADL (+/- SD)** | 7.59 (0.99) | 7.25 (1.29) | 7.95 (0.23)* |
| **ESS (+/- SD)** | 4.59 (3.40) | 3.95 (3.12) | 5.26 (3.63) |
| **ICIQ (+/- SD)** | 2.49 (3.02) | 2.89 (3.43) | 2.06 (2.55) |
| **HADS Depression Score (+/- SD)** | 2.87 (2.30) | 3.95 (2.48) | 1.74 (1.41)* |
| **HADS Anxiety Score (+/- SD)** | 4.38 (2.53) | 4.95 (2.72) | 3.79 (2.23) |
| **QOL-AD – PLWA** | N/A | 37.90 (5.46) | N/A |

| | | | |
|---|---|---|---|
| **QOL-AD - Study Partner** | N/A | 37.23 (4.36) | N/A |
| **Berlin - Low (%)** | 27 (69.23) | 14 (70.00) | 13 (68.42) |
| **Berlin - High** | 12 (30.77) | 6 (30.00) | 6 (31.58) |

Note. PLWA = People Living With Alzheimer's; BMI = Body Mass Index; sMMSE = Standardized Mini-Mental State Examination; PSQI = Pittsburgh Sleep Quality Index. NART = National Adult Reading Test; ADL = Activities of Daily Living; ESS = Epworth Sleepiness Scale; ICIQ = International Consultation on Incontinence Questionnaire; HADS = Hospital Anxiety and Depression Scale; H-O = Horne–Östberg Morningness–Eveningness Questionnaire; AHI = Apnoea–Hypopnea Index; PLMI = Periodic Limb Movement Index; QOL-AD = Quality of Life in Alzheimer's Disease scale. The asterisk symbol indicates significant differences between the PLWA and Control groups.

**Supplemental Table 2.** Mixed model results for biomarkers.

| Biomarker | Group | | Time | | Group*Time | |
|---|---|---|---|---|---|---|
| | *F (DF)* | *P* | *F (DF)* | *P* | *F (DF)* | *P* |
| AB40 | 0.070 (1, 34) | 0.799 | 6.620 (8, 243) | **< .001** | 1.390 (8, 243) | 0.200 |
| AB42 | 1.260 (1, 34) | 0.270 | 8.950 (8, 243) | **< .001** | 2.480 (8, 243) | **0.013** |
| BD-Tau | 3.150 (1, 34) | 0.085 | 4.100 (8, 241) | **< .001** | 0.870 (8, 241) | 0.542 |
| GFAP | 5.170 (1, 34) | **0.029** | 2.740 (8, 243) | **0.007** | 0.790 (8, 243) | 0.614 |
| NfL | 0.140 (1, 34) | 0.715 | 5.030 (8, 243) | **< .001** | 0.420 (8, 243) | 0.907 |
| Cortisol | 0.000 (1, 34) | 0.983 | 46.750 (8, 243) | **< .001** | 0.510 (8, 243) | 0.850 |
| Melatonin | 1.830 (1, 34) | 0.184 | 43.920 (8, 243) | **< .001** | 1.070 (8, 243) | 0.383 |
| p-Tau217 | 20.440 (1, 34) | **< .001** | 4.380 (8, 239) | **< .001** | 2.990 (8, 239) | **0.003** |
| AB42/AB40 | 2.450 (1, 34) | 0.1271 | 2.65 (8, 243) | **0.009** | 1.040 (8, 243) | 0.460 |

**Supplemental Table 3.** Correlations between the mean biomarker levels for all participants.

| Biomarker 1 | Biomarker 2 | r-value | Nominal p-value |
|---|---|---|---|
| AB40 | AB42 | 0.807 | **< .001** |
| GFAP | NfL | 0.729 | **< .001** |
| GFAP | p-Tau217 | 0.705 | **< .001** |
| BD-Tau | GFAP | 0.608 | **< .001** |
| BD-Tau | p-Tau217 | 0.594 | **< .001** |
| AB40 | NfL | 0.57 | **< .001** |
| BD-Tau | AB40 | 0.526 | **< .001** |
| AB42 | AB42/AB40 | 0.517 | **< .001** |
| BD-Tau | NfL | 0.472 | **0.002** |
| NfL | p-Tau217 | 0.418 | **0.008** |
| GFAP | AB42/AB40 | -0.378 | **0.018** |
| AB42 | NfL | 0.372 | **0.02** |
| p-Tau217 | AB42/AB40 | -0.368 | **0.021** |
| GFAP | Cortisol | 0.338 | **0.036** |
| AB40 | GFAP | 0.335 | **0.037** |
| BD-Tau | AB42 | 0.3 | 0.063 |
| NfL | Cortisol | 0.278 | 0.086 |
| BD-Tau | AB42/AB40 | -0.223 | 0.173 |
| AB40 | p-Tau217 | 0.221 | 0.176 |
| AB42 | Melatonin | -0.21 | 0.199 |
| p-Tau217 | Cortisol | 0.202 | 0.218 |
| AB40 | Melatonin | -0.187 | 0.255 |
| GFAP | Melatonin | -0.181 | 0.271 |
| NfL | AB42/AB40 | -0.178 | 0.277 |
| BD-Tau | Cortisol | 0.176 | 0.283 |
| p-Tau217 | Melatonin | -0.169 | 0.303 |
| BD-Tau | Melatonin | -0.161 | 0.327 |
| AB42 | Cortisol | 0.099 | 0.549 |
| NfL | Melatonin | -0.096 | 0.562 |
| AB40 | Cortisol | 0.093 | 0.572 |
| Melatonin | AB42/AB40 | -0.085 | 0.608 |
| AB40 | AB42/AB40 | -0.082 | 0.62 |
| AB42 | GFAP | 0.052 | 0.752 |
| AB42 | p-Tau217 | -0.045 | 0.787 |
| Cortisol | Melatonin | 0.033 | 0.843 |
| Cortisol | AB42/AB40 | 0.024 | 0.883 |

Note. N = 39

**Supplemental Table 4.** Intraclass Correlation Coefficient values for the plasma biomarkers ordered by ICC values.

| Distribution | Biomarker | N | ICC | 95% Lower CI | 95% Upper CI |
|---|---|---|---|---|---|
| *Total sample* | p-Tau217 | 39 | 0.973 | 0.959 | 0.984 |
| | NfL | 39 | 0.956 | 0.932 | 0.974 |
| | AB42/AB40 | 39 | 0.924 | 0.886 | 0.954 |
| | GFAP | 39 | 0.891 | 0.838 | 0.933 |
| | AB42 | 39 | 0.867 | 0.797 | 0.920 |
| | AB40 | 39 | 0.831 | 0.753 | 0.895 |
| | BD-Tau | 39 | 0.782 | 0.692 | 0.861 |
| | Cortisol | 39 | 0.128 | 0.051 | 0.245 |
| | Melatonin | 39 | 0.121 | 0.048 | 0.234 |
| *Controls* | NfL | 19 | 0.966 | 0.939 | 0.985 |
| | p-Tau217 | 19 | 0.952 | 0.912 | 0.978 |
| | AB42/AB40 | 19 | 0.914 | 0.850 | 0.960 |
| | GFAP | 19 | 0.909 | 0.843 | 0.958 |
| | AB42 | 19 | 0.828 | 0.706 | 0.918 |
| | AB40 | 19 | 0.813 | 0.695 | 0.909 |
| | BD-Tau | 19 | 0.710 | 0.560 | 0.849 |
| | Melatonin | 19 | 0.132 | 0.043 | 0.302 |
| | Cortisol | 19 | 0.105 | 0.030 | 0.255 |
| *PLWA* | p-Tau217 | 20 | 0.957 | 0.924 | 0.980 |
| | NfL | 20 | 0.931 | 0.876 | 0.968 |
| | AB42/AB40 | 20 | 0.929 | 0.877 | 0.966 |
| | AB42 | 20 | 0.913 | 0.849 | 0.959 |
| | AB40 | 20 | 0.869 | 0.780 | 0.936 |
| | GFAP | 20 | 0.852 | 0.757 | 0.927 |
| | BD-Tau | 20 | 0.817 | 0.704 | 0.908 |
| | Cortisol | 20 | 0.154 | 0.055 | 0.332 |
| | Melatonin | 20 | 0.110 | 0.033 | 0.261 |

**Supplemental Table 5.** Mixed model results for daytime function.

| **Cognitive measure** | **Group** | | **Study day** | | **Group*Study day** | |
|---|---|---|---|---|---|---|
| | *F (DF)* | *P* | *F (DF)* | *P* | *F (DF)* | *P* |
| Alertness | 2.870 (1, 37.4) | 0.099 | 0.350 (14, 375) | 0.987 | 0.600 (14, 375) | 0.863 |
| Calmness | 2.190 (1, 37.8) | 0.147 | 2.070 (14, 379) | **0.013** | 0.530 (14, 379) | 0.914 |
| Contentedness | 4.650 (1, 37.3) | **0.037** | 0.580 (14, 376) | 0.882 | 0.980 (14, 376) | 0.473 |
| SRT_STD | 2.480 (1, 36.3) | 0.124 | 5.850 (14, 396) | **< .001** | 1.130 (14, 396) | 0.327 |
| SRT_mean | 0.390 (1, 36.5) | 0.536 | 2.120 (14, 390) | **0.010** | 1.610 (14, 390) | 0.073 |
| CRT_STD | 4.490 (1, 38.2) | **0.041** | 2.490 (14, 384) | **0.002** | 0.330 (14, 384) | 0.990 |
| CRT_mean | 1.410 (1, 37.8) | 0.243 | 3.810 (14, 376) | **< .001** | 1.160 (14, 376) | 0.306 |
| Delayed_Recall | 35.950 (1, 37.4) | **< .001** | 2.430 (13, 328) | **0.004** | 1.490 (13, 328) | 0.118 |
| Immediate_Recall | 30.020 (1, 37.5) | **< .001** | 1.540 (14, 389) | 0.095 | 0.490 (14, 389) | 0.938 |
| Evrday_mem_err_tot | 19.700 (1, 36.9) | **< .001** | 3.740 (14, 375) | **< .001** | 0.870 (14, 375) | 0.588 |
| KSS | 4.170 (1, 37.7) | **0.048** | 1.010 (14, 393) | 0.442 | 0.670 (14, 393) | 0.805 |

**Supplemental Table 6.** Correlations between biomarkers and measures of daytime function

| Biomarker | Cognitive Variable | r-value | Nominal p-value |
|---|---|---|---|
| p-Tau217 | Immediate_Recall | -0.604 | **< .001**** |
| p-Tau217 | Delayed_Recall | -0.590 | **< .001**** |
| GFAP | Delayed_Recall | -0.508 | **0.001*** |
| GFAP | Immediate_Recall | -0.484 | **0.002*** |
| BD-Tau | Delayed_Recall | -0.475 | **0.002*** |
| p-Tau217 | Evrday_mem_err_tot | 0.454 | 0.004 |
| p-Tau217 | CRT_STD | 0.424 | 0.007 |
| BD-Tau | CRT_STD | 0.403 | 0.011 |
| BD-Tau | Immediate_Recall | -0.403 | 0.011 |
| NfL | Delayed_Recall | -0.387 | 0.015 |
| GFAP | CRT_STD | 0.379 | 0.017 |
| BD-Tau | CRT_mean | 0.377 | 0.018 |
| Melatonin | SRT_STD | -0.359 | 0.025 |
| p-Tau217 | CRT_mean | 0.320 | 0.047 |
| Melatonin | Evrday_mem_err_tot | -0.314 | 0.052 |
| Melatonin | SRT_mean | -0.298 | 0.065 |
| Melatonin | Contentedness | 0.287 | 0.077 |
| AB42/AB40_ratio | KSS | 0.281 | 0.083 |
| p-Tau217 | SRT_STD | 0.275 | 0.090 |
| NfL | Immediate_Recall | -0.272 | 0.094 |
| GFAP | CRT_mean | 0.272 | 0.094 |
| p-Tau217 | Contentedness | -0.264 | 0.104 |
| GFAP | Evrday_mem_err_tot | 0.259 | 0.112 |
| Cortisol | Delayed_Recall | -0.243 | 0.136 |
| Melatonin | Alertness | 0.238 | 0.144 |
| GFAP | KSS | -0.237 | 0.146 |
| Cortisol | SRT_mean | 0.234 | 0.151 |
| GFAP | SRT_STD | 0.233 | 0.154 |
| Melatonin | Calmness | 0.227 | 0.164 |
| Melatonin | CRT_STD | -0.203 | 0.215 |
| Cortisol | Evrday_mem_err_tot | 0.198 | 0.226 |
| AB42 | Immediate_Recall | 0.195 | 0.233 |
| p-Tau217 | Alertness | -0.195 | 0.235 |
| p-Tau217 | SRT_mean | 0.194 | 0.236 |
| Cortisol | Immediate_Recall | -0.191 | 0.245 |
| NfL | Evrday_mem_err_tot | 0.186 | 0.257 |
| p-Tau217 | Calmness | -0.185 | 0.260 |
| BD-Tau | KSS | -0.182 | 0.268 |
| GFAP | SRT_mean | 0.181 | 0.270 |
| AB42/AB40_ratio | Alertness | -0.174 | 0.290 |
| Cortisol | SRT_STD | 0.171 | 0.297 |
| Melatonin | CRT_mean | -0.170 | 0.302 |
| Cortisol | Contentedness | 0.158 | 0.336 |
| BD-Tau | SRT_STD | 0.157 | 0.339 |
| AB42/AB40_ratio | Immediate_Recall | 0.152 | 0.357 |
| AB42 | Calmness | -0.152 | 0.357 |
| AB40 | KSS | -0.150 | 0.361 |
| NfL | KSS | -0.141 | 0.393 |

| | | | |
|---|---|---|---|
| AB40 | Evrday_mem_err_tot | -0.137 | 0.406 |
| AB42/AB40_ratio | Calmness | -0.135 | 0.412 |
| AB40 | Alertness | 0.134 | 0.416 |
| NfL | SRT_STD | 0.133 | 0.420 |
| AB40 | CRT_mean | 0.128 | 0.438 |
| Cortisol | Alertness | 0.128 | 0.438 |
| AB42/AB40_ratio | Delayed_Recall | 0.127 | 0.440 |
| Cortisol | KSS | -0.127 | 0.441 |
| BD-Tau | SRT_mean | 0.125 | 0.450 |
| AB42/AB40_ratio | Contentedness | -0.124 | 0.451 |
| BD-Tau | Everday_mem_err_tot | 0.122 | 0.460 |
| AB40 | Immediate_Recall | 0.122 | 0.461 |
| AB42/AB40_ratio | SRT_mean | 0.121 | 0.462 |
| p-Tau217 | KSS | 0.121 | 0.462 |
| AB40 | SRT_STD | -0.119 | 0.471 |
| Cortisol | Calmness | 0.118 | 0.473 |
| NfL | SRT_mean | 0.115 | 0.487 |
| NfL | CRT_STD | 0.114 | 0.490 |
| AB42 | Delayed_Recall | 0.108 | 0.512 |
| BD-Tau | Calmness | 0.104 | 0.527 |
| AB42/AB40_ratio | CRT_mean | -0.098 | 0.551 |
| BD-Tau | Contentedness | 0.085 | 0.609 |
| GFAP | Calmness | 0.084 | 0.612 |
| BD-Tau | Alertness | 0.083 | 0.614 |
| Melatonin | Delayed_Recall | 0.081 | 0.626 |
| AB42 | SRT_STD | -0.079 | 0.634 |
| AB42 | Evrday_mem_err_tot | -0.075 | 0.648 |
| AB42/AB40_ratio | Evrday_mem_err_tot | 0.073 | 0.658 |
| NfL | CRT_mean | 0.072 | 0.662 |
| AB40 | Calmness | -0.068 | 0.679 |
| Cortisol | CRT_mean | -0.066 | 0.691 |
| AB42 | SRT_mean | 0.058 | 0.725 |
| Cortisol | CRT_STD | 0.056 | 0.735 |
| AB42 | CRT_mean | 0.055 | 0.740 |
| AB40 | CRT_STD | 0.052 | 0.751 |
| AB42/AB40_ratio | SRT_STD | 0.052 | 0.755 |
| NfL | Contentedness | 0.046 | 0.783 |
| AB40 | Contentedness | 0.043 | 0.797 |
| AB42 | CRT_STD | 0.040 | 0.808 |
| AB42 | Contentedness | -0.037 | 0.822 |
| Melatonin | Immediate_Recall | -0.033 | 0.841 |
| NfL | Calmness | 0.030 | 0.858 |
| AB40 | Delayed_Recall | 0.025 | 0.882 |
| Melatonin | KSS | -0.024 | 0.886 |
| AB42 | KSS | 0.022 | 0.895 |
| AB42 | Alertness | 0.017 | 0.917 |
| GFAP | Alertness | 0.012 | 0.940 |
| GFAP | Contentedness | -0.011 | 0.948 |
| AB42/AB40_ratio | CRT_STD | -0.008 | 0.961 |
| NfL | Alertness | -0.008 | 0.963 |

| | | | |
|---|---|---|---|
| AB40 | SRT_mean | -0.005 | 0.976 |

Note. N = 39. FDR p-value: ** = <0.01, * = <0.05

**Supplemental Table 7.** Correlations between biomarkers and measures of sleep

| Biomarker | WSA Variable | r-value | Nominal p-value |
|---|---|---|---|
| Melatonin | WSA_REMDur | 0.439 | **0.009** |
| AB42 | WSA_DeepSleep_Dur | 0.384 | **0.025** |
| Melatonin | WSA_LightSleepDur | -0.381 | **0.026** |
| AB40 | WSA_DeepSleep_Dur | 0.378 | **0.027** |
| NfL | WSA_TIB | 0.375 | **0.029** |
| AB40 | WSA_TIB | 0.365 | **0.034** |
| Melatonin | WSA_AHI | -0.298 | 0.087 |
| GFAP | WSA_WakeUpLat | -0.289 | 0.097 |
| BD-Tau | WSA_CPD | -0.269 | 0.125 |
| Cortisol | WSA_WakeUpLat | -0.260 | 0.137 |
| NfL | WSA_CPD | -0.252 | 0.151 |
| GFAP | WSA_WASO | 0.249 | 0.155 |
| AB40 | WSA_TST | 0.249 | 0.155 |
| AB42 | WSA_TIB | 0.241 | 0.171 |
| AB42 | WSA_SOL | 0.240 | 0.171 |
| Cortisol | WSA_SOL | 0.234 | 0.183 |
| Melatonin | WSA_CPD | -0.230 | 0.190 |
| BD-Tau | WSA_DeepSleep_Dur | 0.227 | 0.197 |
| NfL | WSA_WakeDur | 0.216 | 0.220 |
| AB42 | WSA_LightSleepDur | -0.211 | 0.230 |
| AB42/AB40_ratio | WSA_LightSleepDur | -0.210 | 0.234 |
| BD-Tau | WSA_WakeUpLat | -0.209 | 0.236 |
| GFAP | WSA_WakeDur | 0.200 | 0.256 |
| NfL | WSA_TST | 0.193 | 0.274 |
| GFAP | WSA_LightSleepDur | 0.192 | 0.278 |
| GFAP | WSA_CPD | -0.190 | 0.283 |
| NfL | WSA_SOL | 0.185 | 0.294 |
| Cortisol | WSA_WakeDur | 0.180 | 0.309 |
| GFAP | WSA_REMDur | -0.179 | 0.312 |
| BD-Tau | p-WSA_REMDur | -0.175 | 0.323 |
| AB42/AB40_ratio | WSA_SOL | 0.170 | 0.336 |
| p-Tau217 | WSA_REMDur | -0.168 | 0.343 |
| NfL | WSA_DeepSleep_Dur | 0.158 | 0.372 |
| GFAP | WSA_SEFF | -0.158 | 0.372 |
| AB40 | WSA_SOL | 0.154 | 0.384 |
| AB42 | WSA_REMDur | -0.150 | 0.396 |
| AB42/AB40_ratio | WSA_TST | -0.149 | 0.399 |
| AB42/AB40_ratio | WSA_REMDur | -0.149 | 0.400 |
| Cortisol | WSA_SEFF | -0.144 | 0.415 |
| Melatonin | WSA_WASO | -0.143 | 0.419 |
| AB42/AB40_ratio | WSA_AHI | 0.141 | 0.426 |

| | | | |
|---|---|---|---|
| GFAP | WSA_AHI | 0.141 | 0.427 |
| AB42 | WSA_WakeDur | 0.137 | 0.441 |
| AB42/AB40_ratio | WSA_WakeUpLat | 0.136 | 0.442 |
| p-Tau217 | WSA_WakeUpLat | -0.135 | 0.445 |
| p-Tau217 | WSA_LightSleepDur | 0.135 | 0.445 |
| p-Tau217 | WSA_SOL | -0.134 | 0.450 |
| Cortisol | WSA_WASO | 0.133 | 0.452 |
| BD-Tau | WSA_TST | 0.127 | 0.474 |
| NfL | WSA_LightSleepDur | 0.126 | 0.478 |
| Melatonin | WSA_DeepSleep_Dur | 0.125 | 0.480 |
| NfL | WSA_WASO | 0.125 | 0.480 |
| AB42 | WSA_TST | 0.125 | 0.482 |
| AB42 | WSA_WakeUpLat | 0.121 | 0.494 |
| AB42/AB40_ratio | WSA_TIB | -0.119 | 0.502 |
| AB42/AB40_ratio | WSA_DeepSleep_Dur | 0.117 | 0.511 |
| NfL | WSA_REMDur | -0.116 | 0.515 |
| AB40 | WSA_LightSleepDur | -0.115 | 0.517 |
| Melatonin | WSA_SOL | 0.112 | 0.529 |
| AB40 | WSA_WakeDur | 0.112 | 0.529 |
| Cortisol | WSA_LightSleepDur | -0.111 | 0.532 |
| AB40 | WSA_AHI | -0.109 | 0.539 |
| BD-Tau | WSA_SEFF | 0.109 | 0.541 |
| Cortisol | WSA_REMDur | 0.108 | 0.544 |
| NfL | WSA_WakeUpLat | -0.105 | 0.555 |
| AB40 | WSA_CPD | -0.103 | 0.562 |
| Cortisol | WSA_CPD | -0.102 | 0.564 |
| BD-Tau | WSA_WASO | -0.102 | 0.566 |
| Melatonin | WSA_TST | 0.100 | 0.572 |
| NfL | WSA_SEFF | -0.100 | 0.573 |
| Melatonin | WSA_WakeDur | -0.100 | 0.574 |
| Cortisol | WSA_TIB | 0.099 | 0.576 |
| AB42/AB40_ratio | WSA_SEFF | -0.099 | 0.576 |
| BD-Tau | WSA_TIB | 0.095 | 0.595 |
| GFAP | WSA_TIB | 0.094 | 0.596 |
| AB42/AB40_ratio | WSA_WASO | 0.088 | 0.619 |
| Cortisol | WSA_AHI | -0.087 | 0.626 |
| p-Tau217 | WSA_TIB | 0.085 | 0.633 |
| GFAP | WSA_DeepSleep_Dur | -0.082 | 0.643 |
| BD-Tau | WSA_SOL | -0.077 | 0.665 |
| Melatonin | WSA_SEFF | 0.074 | 0.678 |
| AB40 | WSA_REMDur | -0.073 | 0.682 |
| AB42/AB40_ratio | WSA_WakeDur | 0.072 | 0.687 |
| AB42 | WSA_SEFF | -0.071 | 0.691 |

| | | | |
|---|---|---|---|
| BD-Tau | WSA_WakeDur | -0.070 | 0.692 |
| p-Tau217 | WSA_AHI | 0.069 | 0.700 |
| p-Tau217 | WSA_TST | 0.063 | 0.722 |
| AB42 | WSA_WASO | 0.060 | 0.735 |
| p-Tau217 | WSA_DeepSleep_Dur | 0.058 | 0.744 |
| AB42/AB40_ratio | WSA_CPD | 0.052 | 0.771 |
| GFAP | WSA_TST | -0.043 | 0.809 |
| Melatonin | WSA_TIB | 0.043 | 0.810 |
| p-Tau217 | WSA_SEFF | 0.041 | 0.818 |
| GFAP | WSA_SOL | 0.038 | 0.829 |
| AB42 | WSA_CPD | -0.035 | 0.844 |
| Cortisol | WSA_TST | -0.026 | 0.884 |
| NfL | WSA_AHI | 0.023 | 0.896 |
| p-Tau217 | WSA_WakeDur | 0.018 | 0.921 |
| BD-Tau | WSA_AHI | 0.017 | 0.926 |
| AB40 | WSA_SEFF | -0.014 | 0.936 |
| p-Tau217 | WSA_WASO | 0.011 | 0.950 |
| AB40 | WSA_WakeUpLat | 0.011 | 0.952 |
| AB42 | WSA_AHI | 0.011 | 0.952 |
| AB40 | WSA_WASO | 0.010 | 0.956 |
| p-Tau217 | WSA_CPD | -0.007 | 0.969 |
| BD-Tau | WSA_LightSleepDur | 0.007 | 0.971 |
| Cortisol | WSA_DeepSleep_Dur | -0.004 | 0.982 |
| Melatonin | WSA_WakeUpLat | -0.001 | 0.996 |

Note. N = 34. FDR p-value: N.S.

**Supplemental Table 8.** Mixed model results for correlation magnitude between daytime function and biomarker levels across the 24-hour day.

| Biomarker (DV) | Cognitive measure list | Time | | Cognitive measure | | Time*Cognitive measure | |
|---|---|---|---|---|---|---|---|
| | | *F (DF)* | *P* | *F (DF)* | *P* | *F (DF)* | *P* |
| AB40 | Alertness | 7.214 (8, 259.026) | **< .001** | 0.822 (1, 36.778) | 0.371 | 0.765 (8, 259.003) | 0.634 |
| | CRT_STD | 0.851 (8, 259.183) | 0.558 | 0.071 (1, 37.035) | 0.791 | 0.909 (8, 259.193) | 0.510 |
| | CRT_mean | 0.445 (8, 259.272) | 0.893 | 0.491 (1, 37.120) | 0.488 | 0.455 (8, 259.276) | 0.886 |
| | Calmness | 7.397 (8, 259.094) | **< .001** | 0.104 (1, 36.991) | 0.749 | 1.296 (8, 259.191) | 0.246 |
| | Contentedness | 7.410 (8, 259.048) | **< .001** | 0.122 (1, 36.929) | 0.728 | 1.289 (8, 259.125) | 0.249 |
| | Delayed_Recall | 0.564 (8, 259.120) | 0.807 | 0.032 (1, 36.970) | 0.858 | 0.564 (8, 259.110) | 0.807 |
| | Evrday_mem_err_tot | 1.819 (8, 258.969) | 0.074 | 0.767 (1, 36.821) | 0.387 | 0.627 (8, 259.041) | 0.755 |
| | Immediate_Recall | 0.816 (8, 259.022) | 0.589 | 0.643 (1, 36.880) | 0.428 | 0.831 (8, 259.018) | 0.576 |
| | KSS | 2.010 (8, 258.947) | **0.046** | 0.914 (1, 36.726) | 0.345 | 1.282 (8, 258.952) | 0.253 |
| | SRT_STD | 1.599 (8, 259.045) | 0.125 | 0.509 (1, 36.829) | 0.480 | 1.783 (8, 259.054) | 0.081 |
| | SRT_mean | 0.586 (8, 259.056) | 0.790 | 0.000 (1, 36.868) | 0.989 | 0.575 (8, 259.060) | 0.798 |
| AB42 | Alertness | 10.497 (8, 259.079) | **< .001** | 0.029 (1, 36.911) | 0.864 | 0.449 (8, 259.063) | 0.891 |
| | CRT_STD | 0.786 (8, 259.164) | 0.615 | 0.043 (1, 37.062) | 0.837 | 0.737 (8, 259.171) | 0.659 |
| | CRT_mean | 0.531 (8, 259.201) | 0.833 | 0.072 (1, 37.096) | 0.790 | 0.515 (8, 259.204) | 0.845 |
| | Calmness | 10.715 (8, 259.119) | **< .001** | 0.714 (1, 37.046) | 0.404 | 0.872 (8, 259.189) | 0.540 |
| | Contentedness | 10.797 (8, 259.094) | **< .001** | 0.024 (1, 37.011) | 0.878 | 0.949 (8, 259.149) | 0.476 |
| | Delayed_Recall | 0.983 (8, 259.091) | 0.450 | 0.457 (1, 36.962) | 0.503 | 1.170 (8, 259.084) | 0.318 |
| | Evrday_mem_err_tot | 3.950 (8, 259.028) | **< .001** | 0.233 (1, 36.935) | 0.632 | 1.013 (8, 259.078) | 0.427 |
| | Immediate_Recall | 1.381 (8, 259.025) | 0.205 | 1.540 (1, 36.926) | 0.222 | 1.569 (8, 259.022) | 0.134 |
| | KSS | 2.407 (8, 259.030) | **0.016** | 0.011 (1, 36.874) | 0.917 | 1.288 (8, 259.034) | 0.250 |
| | SRT_STD | 1.509 (8, 259.070) | 0.154 | 0.216 (1, 36.917) | 0.645 | 1.500 (8, 259.077) | 0.157 |
| | SRT_mean | 0.913 (8, 259.077) | 0.506 | 0.151 (1, 36.927) | 0.700 | 0.876 (8, 259.080) | 0.538 |
| BD-Tau | Alertness | 4.815 (8, 257.276) | **< .001** | 0.272 (1, 36.941) | 0.605 | 1.525 (8, 257.272) | 0.149 |
| | CRT_STD | 1.449 (8, 257.739) | 0.176 | 7.603 (1, 37.419) | **0.009** | 1.618 (8, 257.759) | 0.120 |
| | CRT_mean | 3.545 (8, 257.806) | **0.001** | 6.437 (1, 37.521) | **0.016** | 3.610 (8, 257.814) | **< .001** |
| | Calmness | 4.942 (8, 257.278) | **< .001** | 0.441 (1, 37.159) | 0.511 | 1.161 (8, 257.478) | 0.323 |
| | Contentedness | 4.932 (8, 257.280) | **< .001** | 0.267 (1, 37.111) | 0.609 | 1.080 (8, 257.430) | 0.378 |

| | | | | | | | |
|---|---|---|---|---|---|---|---|
| | Delayed_Recall | 0.854 (8, 257.407) | 0.556 | 10.810 (1, 37.050) | **0.002** | 0.765 (8, 257.390) | 0.634 |
| | Evrday_mem_err_tot | 1.122 (8, 257.269) | 0.349 | 0.611 (1, 37.003) | 0.440 | 0.874 (8, 257.352) | 0.539 |
| | Immediate_Recall | 2.046 (8, 257.406) | **0.042** | 6.771 (1, 37.153) | **0.013** | 1.920 (8, 257.399) | 0.057 |
| | KSS | 1.584 (8, 257.197) | 0.130 | 1.266 (1, 36.812) | 0.268 | 0.955 (8, 257.193) | 0.472 |
| | SRT_STD | 0.987 (8, 257.291) | 0.446 | 1.107 (1, 36.930) | 0.300 | 1.268 (8, 257.310) | 0.261 |
| | SRT_mean | 1.229 (8, 257.278) | 0.282 | 0.651 (1, 36.913) | 0.425 | 1.192 (8, 257.285) | 0.304 |
| GFAP | Alertness | 4.198 (8, 259.099) | **< .001** | 0.009 (1, 36.984) | 0.926 | 1.316 (8, 259.084) | 0.236 |
| | CRT_STD | 0.987 (8, 259.256) | 0.447 | 6.158 (1, 37.153) | **0.018** | 1.136 (8, 259.264) | 0.340 |
| | CRT_mean | 0.997 (8, 259.252) | 0.439 | 2.930 (1, 37.149) | 0.095 | 1.046 (8, 259.255) | 0.402 |
| | Calmness | 4.282 (8, 259.075) | **< .001** | 0.281 (1, 37.008) | 0.599 | 0.373 (8, 259.141) | 0.934 |
| | Contentedness | 4.407 (8, 259.101) | **< .001** | 0.003 (1, 37.023) | 0.959 | 0.817 (8, 259.152) | 0.588 |
| | Delayed_Recall | 1.554 (8, 259.197) | 0.139 | 12.516 (1, 37.040) | **0.001** | 1.389 (8, 259.188) | 0.202 |
| | Evrday_mem_err_tot | 0.418 (8, 259.123) | 0.910 | 2.647 (1, 37.025) | 0.112 | 1.220 (8, 259.172) | 0.288 |
| | Immediate_Recall | 1.067 (8, 259.135) | 0.386 | 10.973 (1, 37.049) | **0.002** | 0.942 (8, 259.131) | 0.482 |
| | KSS | 1.191 (8, 259.028) | 0.305 | 2.250 (1, 36.908) | 0.142 | 0.747 (8, 259.032) | 0.650 |
| | SRT_STD | 2.604 (8, 259.121) | **0.009** | 2.134 (1, 36.976) | 0.153 | 2.995 (8, 259.127) | **0.003** |
| | SRT_mean | 1.841 (8, 259.117) | 0.070 | 1.263 (1, 36.976) | 0.268 | 1.905 (8, 259.120) | 0.060 |
| NfL | Alertness | 6.856 (8, 259.034) | **< .001** | 0.001 (1, 36.990) | 0.974 | 2.083 (8, 259.029) | **0.038** |
| | CRT_STD | 1.249 (8, 259.064) | 0.271 | 0.488 (1, 37.033) | 0.489 | 1.462 (8, 259.067) | 0.172 |
| | CRT_mean | 0.621 (8, 259.075) | 0.760 | 0.190 (1, 37.041) | 0.666 | 0.666 (8, 259.076) | 0.721 |
| | Calmness | 6.669 (8, 259.030) | **< .001** | 0.037 (1, 37.025) | 0.848 | 0.204 (8, 259.054) | 0.990 |
| | Contentedness | 6.867 (8, 259.028) | **< .001** | 0.083 (1, 37.009) | 0.775 | 0.698 (8, 259.047) | 0.693 |
| | Delayed_Recall | 1.321 (8, 259.076) | 0.233 | 6.398 (1, 37.031) | **0.016** | 1.123 (8, 259.073) | 0.348 |
| | Evrday_mem_err_tot | 0.809 (8, 259.042) | 0.595 | 1.322 (1, 37.012) | 0.258 | 1.098 (8, 259.059) | 0.365 |
| | Immediate_Recall | 0.943 (8, 259.041) | 0.482 | 2.822 (1, 37.033) | 0.101 | 0.784 (8, 259.040) | 0.617 |
| | KSS | 1.108 (8, 259.010) | 0.358 | 0.762 (1, 36.988) | 0.388 | 0.617 (8, 259.011) | 0.763 |
| | SRT_STD | 2.204 (8, 259.039) | **0.028** | 0.691 (1, 36.988) | 0.411 | 2.681 (8, 259.041) | **0.008** |
| | SRT_mean | 0.883 (8, 259.037) | 0.531 | 0.512 (1, 36.987) | 0.479 | 0.868 (8, 259.038) | 0.544 |
| p-Tau217 | Alertness | 4.752 (8, 252.987) | **< .001** | 1.442 (1, 36.947) | 0.238 | 1.721 (8, 252.984) | 0.094 |

| | | | | | | | |
|---|---|---|---|---|---|---|---|
| | CRT_STD | 1.131 (8, 253.009) | 0.343 | 8.270 (1, 36.981) | **0.007** | 1.240 (8, 253.011) | 0.276 |
| | CRT_mean | 0.687 (8, 253.012) | 0.703 | 4.285 (1, 36.980) | **0.046** | 0.716 (8, 253.012) | 0.678 |
| | Calmness | 4.753 (8, 252.988) | **< .001** | 1.296 (1, 36.966) | 0.262 | 0.962 (8, 253.001) | 0.466 |
| | Contentedness | 4.855 (8, 252.993) | **< .001** | 2.796 (1, 36.972) | 0.103 | 1.422 (8, 253.006) | 0.187 |
| | Delayed_Recall | 3.057 (8, 252.983) | **0.003** | 19.639 (1, 36.936) | **< .001** | 2.821 (8, 252.981) | **0.005** |
| | Evrday_mem_err_tot | 1.613 (8, 252.980) | 0.121 | 9.744 (1, 36.950) | **0.004** | 2.786 (8, 252.994) | **0.006** |
| | Immediate_Recall | 2.854 (8, 252.950) | **0.005** | 21.041 (1, 36.914) | **< .001** | 2.631 (8, 252.949) | **0.009** |
| | KSS | 0.634 (8, 252.975) | 0.749 | 0.538 (1, 36.937) | 0.468 | 0.767 (8, 252.976) | 0.632 |
| | SRT_STD | 1.348 (8, 252.990) | 0.220 | 3.121 (1, 36.947) | 0.086 | 1.564 (8, 252.992) | 0.136 |
| | SRT_mean | 0.747 (8, 252.985) | 0.650 | 1.497 (1, 36.946) | 0.229 | 0.787 (8, 252.986) | 0.614 |
| AB42/ AB40_ratio | Alertness | 3.518 (8, 259.133) | **0.001** | 1.154 (1, 37.020) | 0.290 | 1.029 (8, 259.123) | 0.415 |
| | CRT_STD | 0.888 (8, 259.176) | 0.527 | 0.000 (1, 37.110) | 0.983 | 0.839 (8, 259.180) | 0.569 |
| | CRT_mean | 0.494 (8, 259.209) | 0.860 | 0.345 (1, 37.141) | 0.560 | 0.473 (8, 259.210) | 0.875 |
| | Calmness | 3.501 (8, 259.131) | **0.001** | 0.674 (1, 37.085) | 0.417 | 1.301 (8, 259.176) | 0.243 |
| | Contentedness | 3.464 (8, 259.129) | **0.001** | 0.577 (1, 37.075) | 0.452 | 1152 (8, 259.165) | 0.329 |
| | Delayed_Recall | 2.306 (8, 259.138) | **0.021** | 0.616 (1, 37.096) | 0.437 | 2.460 (8, 259.133) | **0.014** |
| | Evrday_mem_err_tot | 2.722 (8, 259.115) | **0.007** | 0.203 (1, 37.053) | 0.655 | 1.091 (8, 259.148) | 0.370 |
| | Immediate_Recall | 1.880 (8, 259.102) | 0.063 | 0.843 (1, 37.049) | 0.365 | 2.010 (8, 259.100) | **0.046** |
| | KSS | 0.640 (8, 259.103) | 0.744 | 3.141 (1, 37.003) | 0.085 | 0.620 (8, 259.106) | 0.761 |
| | SRT_STD | 1.911 (8, 259.128) | 0.059 | 0.103 (1, 37.028) | 0.750 | 1.711 (8, 259.133) | 0.096 |
| | SRT_mean | 0.839 (8, 259.122) | 0.569 | 0.600 (1, 37.070) | 0.443 | 0.810 (8, 259.124) | 0.594 |
| Melatonin | Alertness | 57.446 (8, 261.238) | 0.000 | 4.316 (1, 37.567) | 0.045 | 3.994 (8, 261.066) | 0.000 |
| | CRT_STD | 1.964 (8, 262.229) | 0.051 | 1.046 (1, 39.381) | 0.313 | 1.332 (8, 262.314) | 0.228 |
| | CRT_mean | 2.366 (8, 262.942) | 0.018 | 1.306 (1, 40.113) | 0.260 | 2.068 (8, 262.978) | 0.039 |
| | Calmness | 55.499 (8, 261.256) | 0.000 | 3.885 (1, 38.598) | 0.056 | 3.127 (8, 261.945) | 0.002 |
| | Contentedness | 56.661 (8, 261.381) | 0.000 | 5.632 (1, 38.554) | 0.023 | 4.082 (8, 261.933) | 0.000 |
| | Delayed_Recall | 0.271 (8, 262.505) | 0.975 | 0.173 (1, 39.542) | 0.680 | 0.303 (8, 262.399) | 0.965 |
| | Evrday_mem_err_tot | 25.201 (8, 261.019) | 0.000 | 3.674 (1, 38.153) | 0.063 | 3.090 (8, 261.685) | 0.002 |
| | Immediate_Recall | 0.682 (8, 261.467) | 0.707 | 0.009 (1, 38.962) | 0.927 | 0.356 (8, 261.415) | 0.942 |
| | KSS | 2.743 (8, 260.711) | 0.006 | 0.117 (1, 37.115) | 0.734 | 0.697 (8, 260.742) | 0.694 |

| | | | | | | | |
|---|---|---|---|---|---|---|---|
| | SRT_STD | 5.245 (8, 262.007) | 0.000 | 4.353 (1, 38.458) | 0.044 | 3.226 (8, 262.136) | 0.002 |
| | SRT_mean | 1.901 (8, 261.877) | 0.060 | 1.875 (1, 38.300) | 0.179 | 1.445 (8, 261.924) | 0.178 |
| Cortisol | Alertness | 58.319 (8, 260.327) | 0.000 | 0.166 (1, 36.796) | 0.686 | 1.248 (8, 260.160) | 0.272 |
| | CRT_STD | 1.711 (8, 260.957) | 0.096 | 0.112 (1, 38.185) | 0.739 | 1.301 (8, 261.039) | 0.243 |
| | CRT_mean | 0.758 (8, 261.692) | 0.640 | 0.046 (1, 38.861) | 0.831 | 0.625 (8, 261.727) | 0.756 |
| | Calmness | 57.110 (8, 260.378) | 0.000 | 0.344 (1, 37.767) | 0.561 | 0.664 (8, 261.052) | 0.723 |
| | Contentedness | 57.007 (8, 260.351) | 0.000 | 0.815 (1, 37.568) | 0.372 | 0.632 (8, 260.895) | 0.751 |
| | Delayed_Recall | 1.399 (8, 261.859) | 0.197 | 3.838 (1, 38.807) | 0.057 | 1.163 (8, 261.752) | 0.322 |
| | Evrday_mem_err_tot | 15.941 (8, 260.371) | 0.000 | 1.335 (1, 37.566) | 0.255 | 1.001 (8, 261.022) | 0.436 |
| | Immediate_Recall | 1.362 (8, 260.731) | 0.213 | 1.930 (1, 38.220) | 0.173 | 1.145 (8, 260.680) | 0.334 |
| | KSS | 2.444 (8, 259.942) | 0.014 | 0.428 (1, 36.454) | 0.517 | 1.333 (8, 259.973) | 0.227 |
| | SRT_STD | 1.992 (8, 260.761) | 0.048 | 0.883 (1, 37.451) | 0.353 | 1.471 (8, 260.880) | 0.168 |
| | SRT_mean | 0.907 (8, 260.415) | 0.511 | 1.336 (1, 36.955) | 0.255 | 0.892 (8, 260.460) | 0.524 |

**Supplemental Table 9.** Intraclass Correlation Coefficient values for the daytime function measures ordered by ICC value.

| Distribution | Measure | N | ICC | 95% Lower CI | 95% Upper CI |
|---|---|---|---|---|---|
| Total sample | *Everday_mem_err_tot* | 39 | 0.862 | 0.801 | 0.913 |
| | *Alertness* | 39 | 0.809 | 0.734 | 0.878 |
| | *Contentedness* | 39 | 0.803 | 0.726 | 0.873 |
| | *SRT_mean* | 39 | 0.761 | 0.674 | 0.844 |
| | *CRT_mean* | 39 | 0.710 | 0.610 | 0.807 |
| | *Calmness* | 39 | 0.698 | 0.599 | 0.797 |
| | *Immediate_Recall* | 39 | 0.535 | 0.423 | 0.665 |
| | *CRT_STD* | 39 | 0.519 | 0.406 | 0.650 |
| | *Delayed_Recall* | 39 | 0.514 | 0.401 | 0.645 |
| | *KSS* | 39 | 0.491 | 0.378 | 0.625 |
| | *SRT_STD* | 39 | 0.462 | 0.350 | 0.599 |
| Controls | *Everday_mem_err_tot* | 19 | 0.869 | 0.785 | 0.937 |
| | *Alertness* | 19 | 0.801 | 0.688 | 0.900 |
| | *Contentedness* | 19 | 0.788 | 0.670 | 0.893 |
| | *SRT_mean* | 19 | 0.779 | 0.658 | 0.887 |
| | *CRT_mean* | 19 | 0.745 | 0.614 | 0.868 |
| | *Calmness* | 19 | 0.676 | 0.531 | 0.825 |
| | *CRT_STD* | 19 | 0.603 | 0.448 | 0.775 |
| | *KSS* | 19 | 0.524 | 0.367 | 0.716 |
| | *SRT_STD* | 19 | 0.488 | 0.333 | 0.687 |
| | *Delayed_Recall* | 19 | 0.388 | 0.244 | 0.597 |
| | *Immediate_Recall* | 19 | 0.358 | 0.219 | 0.568 |
| PLWA | *Alertness* | 20 | 0.809 | 0.702 | 0.903 |
| | *Contentedness* | 20 | 0.792 | 0.679 | 0.893 |
| | *SRT_mean* | 20 | 0.746 | 0.618 | 0.866 |
| | *Calmness* | 20 | 0.707 | 0.571 | 0.841 |

| | | | | |
|---|---|---|---|---|
| *Everday_mem_err_tot* | 20 | 0.678 | 0.536 | 0.823 |
| *CRT_mean* | 20 | 0.656 | 0.507 | 0.808 |
| *KSS* | 20 | 0.408 | 0.264 | 0.611 |
| *SRT_STD* | 20 | 0.404 | 0.260 | 0.606 |
| *Immediate_Recall* | 20 | 0.391 | 0.249 | 0.594 |
| *CRT_STD* | 20 | 0.298 | 0.174 | 0.497 |
| *Delayed_Recall* | 20 | 0.289 | 0.167 | 0.488 |

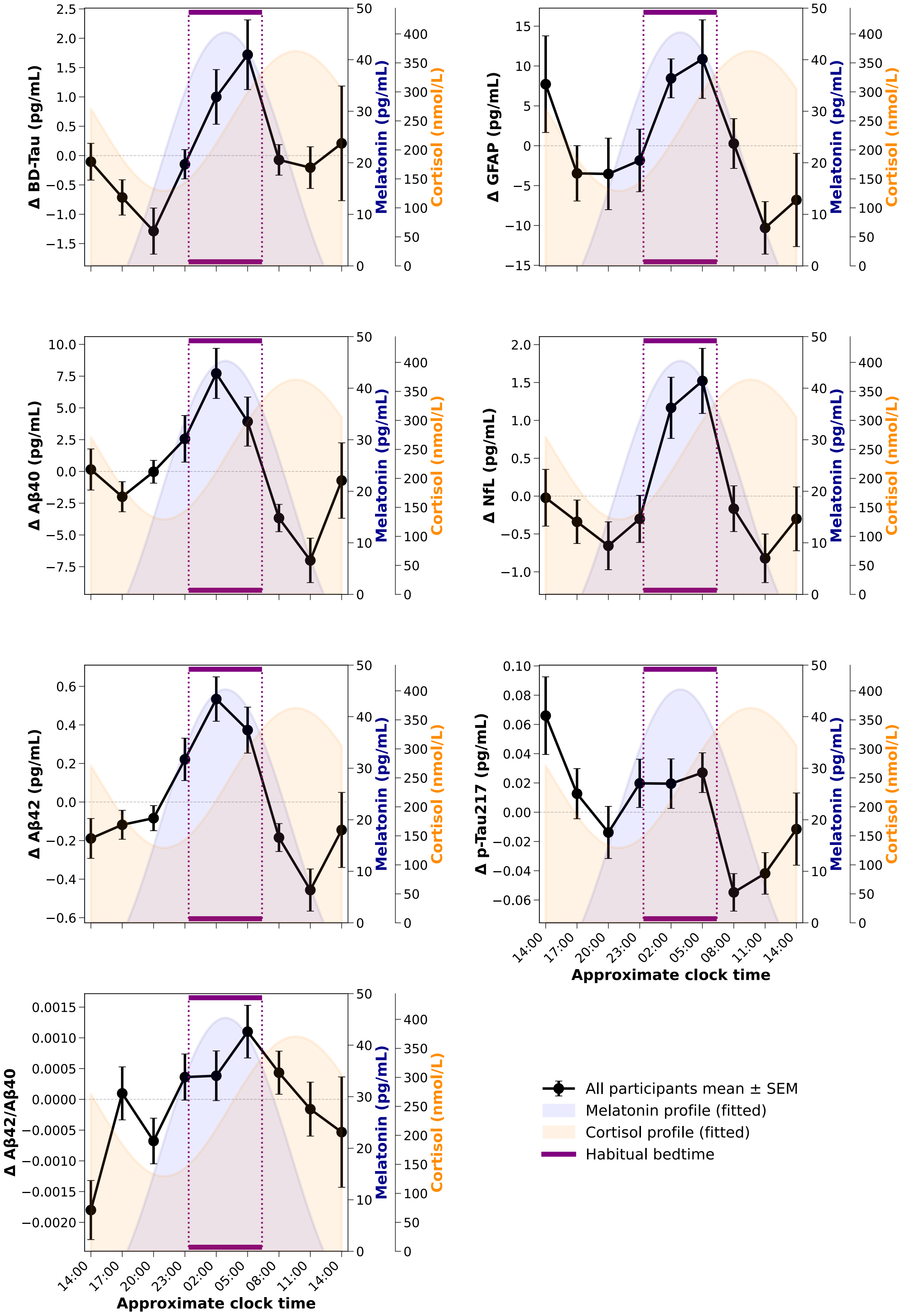
Δ BD-Tau (pg/mL)
Δ GFAP (pg/mL)
Δ Aβ40 (pg/mL)
Δ NfL (pg/mL)
Δ Aβ42 (pg/mL)
Δ p-Tau217 (pg/mL)
Δ Aβ42/Aβ40
Melatonin (pg/mL)
Cortisol (nmol/L)
14:00
17:00
20:00
23:00
02:00
05:00
08:00
11:00
14:00
Approximate clock time
All participants mean ± SEM
Melatonin profile (fitted)
Cortisol profile (fitted)
Habitual bedtime

**Supplemental Figure 1.** Deviation from the mean and SE for the 24-hour profiles of blood-based biomarkers of dementia for all participants (black lines) superimposed on average melatonin (blue shading) and cortisol (yellow shading) profiles. Average habitual bedtime is indicated by the purple horizontal lines. The deviation from the mean values were calculated per participant and then the mean was calculated for all participants.

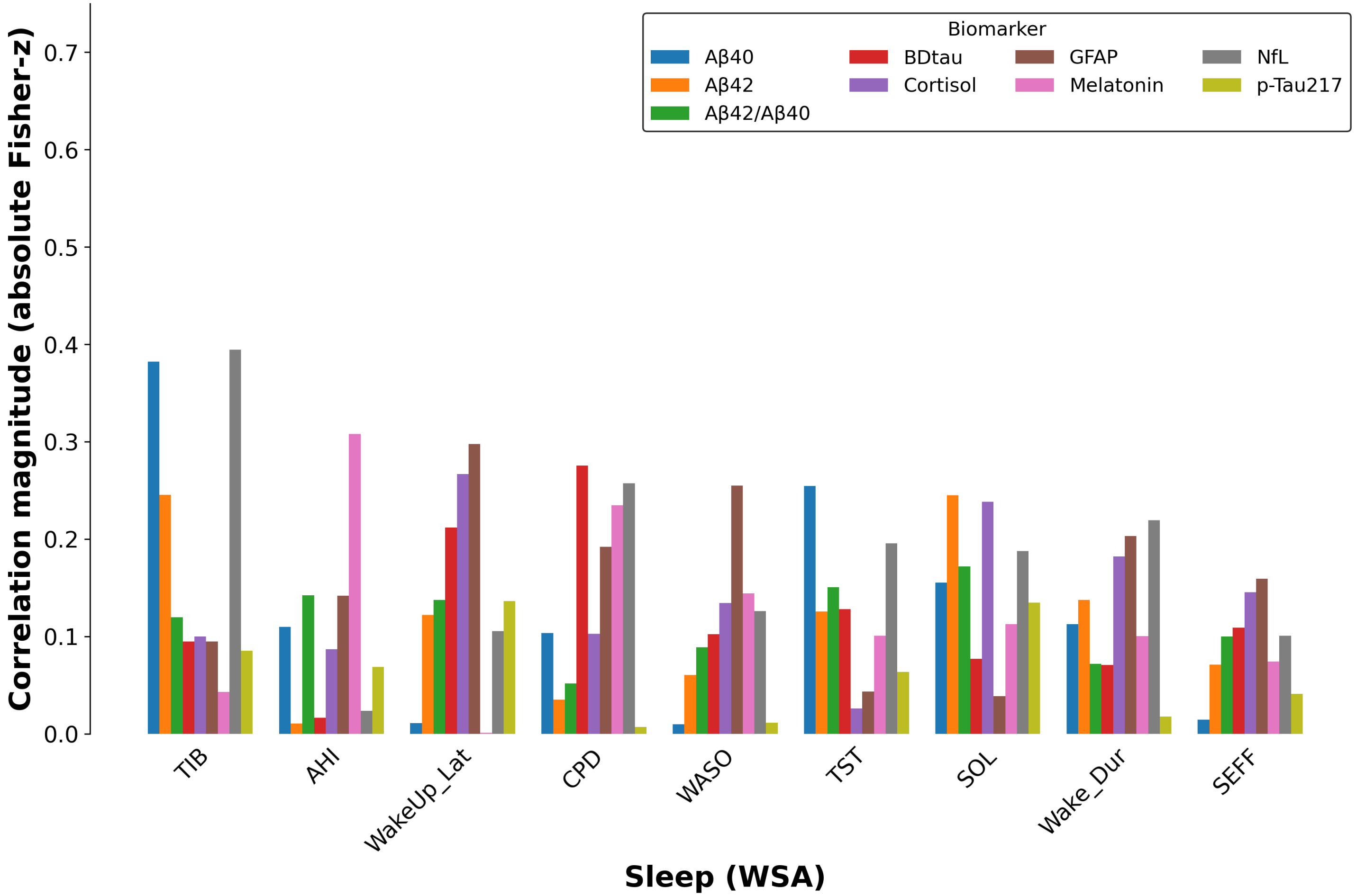


**Supplemental Figure 2.** Correlation magnitudes between levels (mean of nine samples) of eight blood-based biomarkers and measures of sleep (mean of 15 nights).

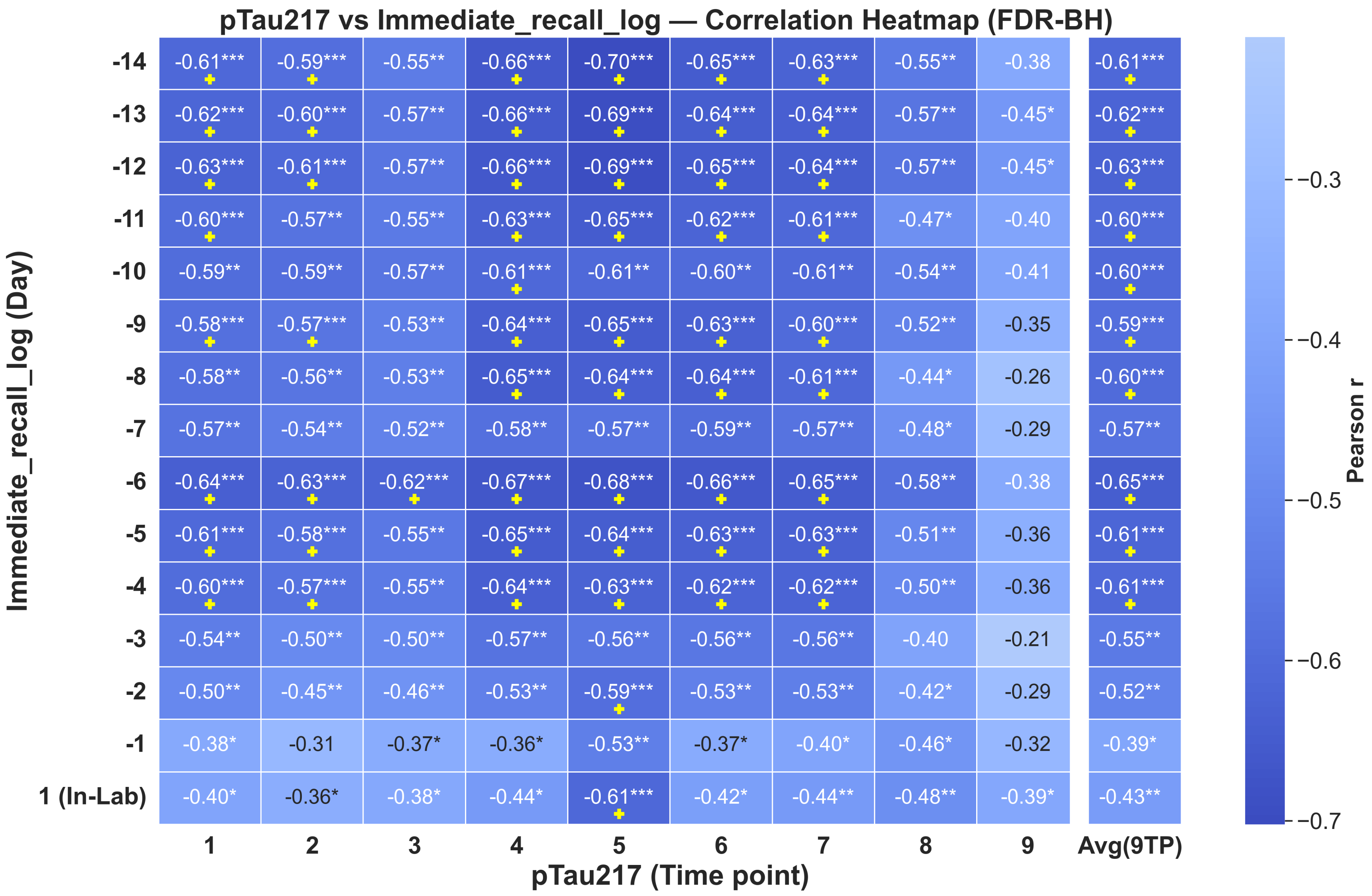
pTau217 vs Immediate_recall_log — Correlation Heatmap (FDR-BH)
Immediate_recall_log (Day)
pTau217 (Time point)
1
2
3
4
5
6
7
8
9
Avg(9TP)
-14 -0.61*** -0.59*** -0.55** -0.66*** -0.70*** -0.65*** -0.63*** -0.55** -0.38 -0.61***
-13 -0.62*** -0.60*** -0.57** -0.66*** -0.69*** -0.64*** -0.64*** -0.57** -0.45* -0.62***
-12 -0.63*** -0.61*** -0.57** -0.66*** -0.69*** -0.65*** -0.64*** -0.57** -0.45* -0.63***
-11 -0.60*** -0.57** -0.55** -0.63*** -0.65*** -0.62*** -0.61*** -0.47* -0.40 -0.60***
-10 -0.59** -0.59** -0.57** -0.61*** -0.61** -0.60** -0.61** -0.54** -0.41 -0.60***
-9 -0.58*** -0.57*** -0.53** -0.64*** -0.65*** -0.63*** -0.60*** -0.52** -0.35 -0.59***
-8 -0.58** -0.56** -0.53** -0.65*** -0.64*** -0.64*** -0.61*** -0.44* -0.26 -0.60***
-7 -0.57** -0.54** -0.52** -0.58** -0.57** -0.59** -0.57** -0.48* -0.29 -0.57**
-6 -0.64*** -0.63*** -0.62*** -0.67*** -0.68*** -0.66*** -0.65*** -0.58** -0.38 -0.65***
-5 -0.61*** -0.58*** -0.55** -0.65*** -0.64*** -0.63*** -0.63*** -0.51** -0.36 -0.61***
-4 -0.60*** -0.57*** -0.55** -0.64*** -0.63*** -0.62*** -0.62*** -0.50** -0.36 -0.61***
-3 -0.54** -0.50** -0.50** -0.57** -0.56** -0.56** -0.56** -0.40 -0.21 -0.55**
-2 -0.50** -0.45** -0.46** -0.53** -0.59*** -0.53** -0.53** -0.42* -0.29 -0.52**
-1 -0.38* -0.31 -0.37* -0.36* -0.53** -0.37* -0.40* -0.46* -0.32 -0.39*
1 (In-Lab) -0.40* -0.36* -0.38* -0.44* -0.61*** -0.42* -0.44** -0.48** -0.39* -0.43**
Pearson r
−0.3
−0.4
−0.5
−0.6
−0.7

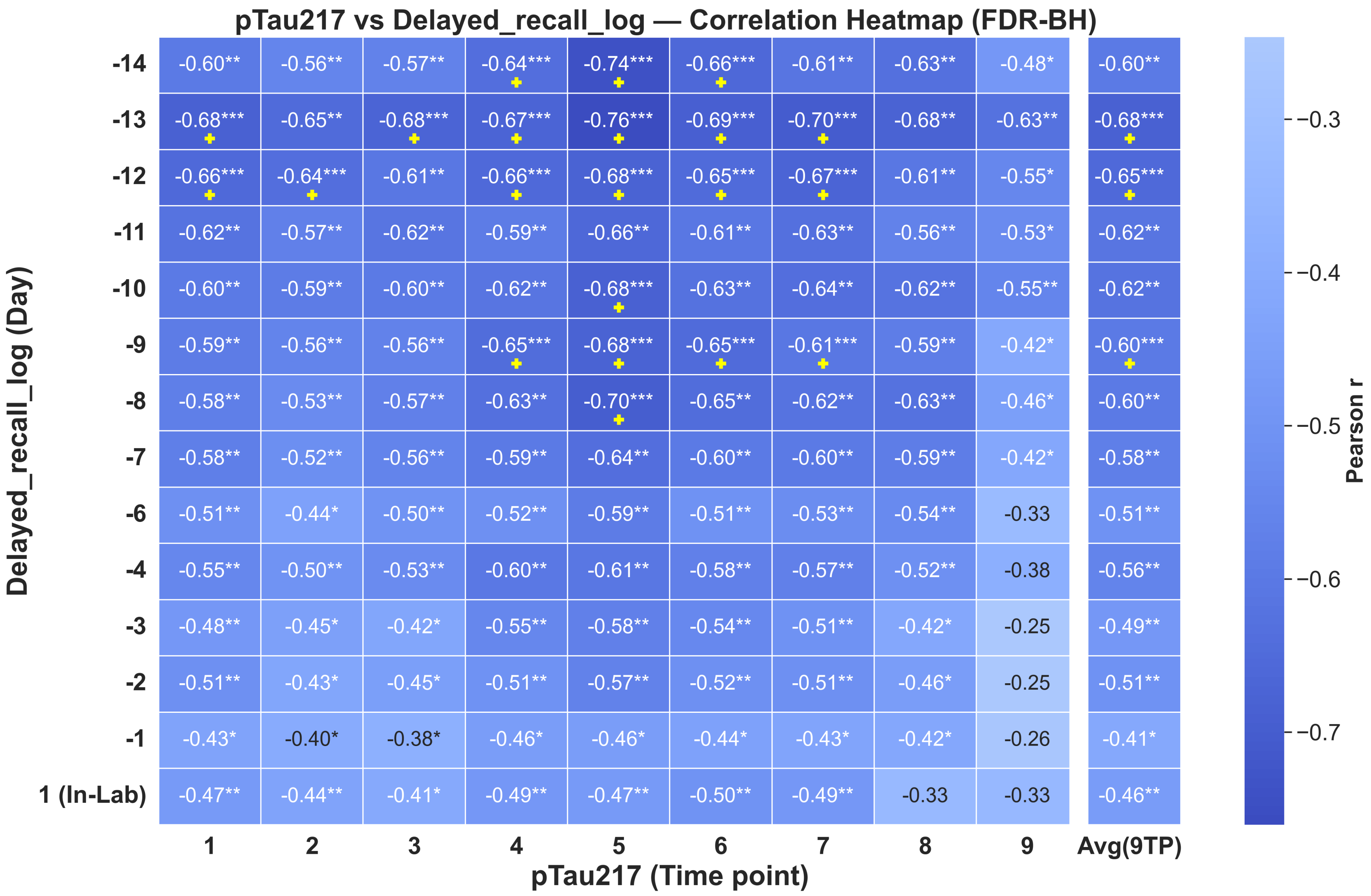
pTau217 vs Delayed_recall_log — Correlation Heatmap (FDR-BH)
Delayed_recall_log (Day)
pTau217 (Time point)
1 2 3 4 5 6 7 8 9 Avg(9TP)
-14 -0.60** -0.56** -0.57** -0.64*** -0.74*** -0.66*** -0.61** -0.63** -0.48* -0.60**
-13 -0.68*** -0.65** -0.68*** -0.67*** -0.76*** -0.69*** -0.70*** -0.68** -0.63** -0.68***
-12 -0.66*** -0.64*** -0.61** -0.66*** -0.68*** -0.65*** -0.67*** -0.61** -0.55* -0.65***
-11 -0.62** -0.57** -0.62** -0.59** -0.66** -0.61** -0.63** -0.56** -0.53* -0.62**
-10 -0.60** -0.59** -0.60** -0.62** -0.68*** -0.63** -0.64** -0.62** -0.55** -0.62**
-9 -0.59** -0.56** -0.56** -0.65*** -0.68*** -0.65*** -0.61*** -0.59** -0.42* -0.60***
-8 -0.58** -0.53** -0.57** -0.63** -0.70*** -0.65** -0.62** -0.63** -0.46* -0.60**
-7 -0.58** -0.52** -0.56** -0.59** -0.64** -0.60** -0.60** -0.59** -0.42* -0.58**
-6 -0.51** -0.44* -0.50** -0.52** -0.59** -0.51** -0.53** -0.54** -0.33 -0.51**
-4 -0.55** -0.50** -0.53** -0.60** -0.61** -0.58** -0.57** -0.52** -0.38 -0.56**
-3 -0.48** -0.45* -0.42* -0.55** -0.58** -0.54** -0.51** -0.42* -0.25 -0.49**
-2 -0.51** -0.43* -0.45* -0.51** -0.57** -0.52** -0.51** -0.46* -0.25 -0.51**
-1 -0.43* -0.40* -0.38* -0.46* -0.46* -0.44* -0.43* -0.42* -0.26 -0.41*
1 (In-Lab) -0.47** -0.44** -0.41* -0.49** -0.47** -0.50** -0.49** -0.33 -0.33 -0.46**
Pearson r
−0.3
−0.4
−0.5
−0.6
−0.7

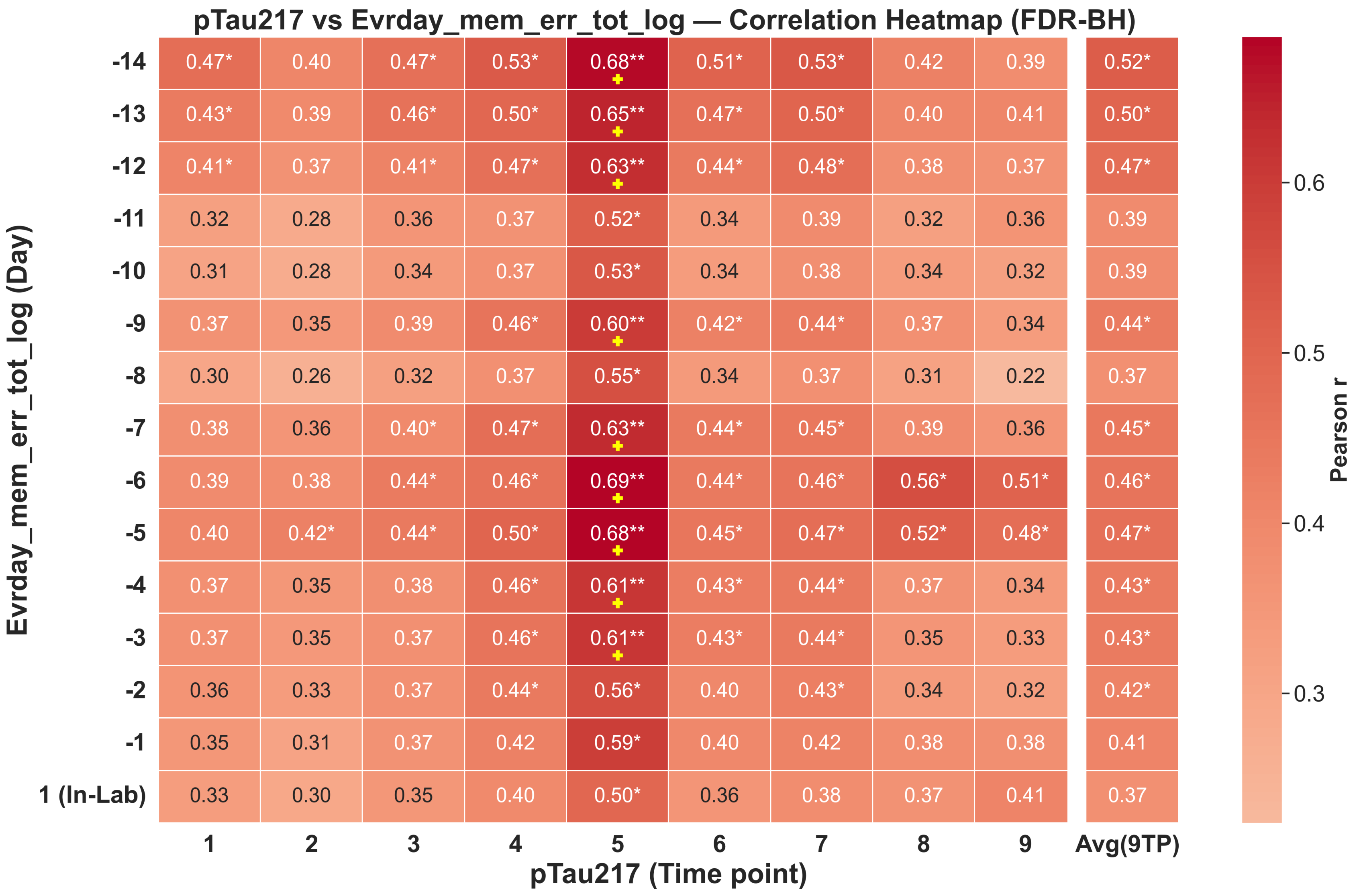

pTau217 vs Evrday_mem_err_tot_log — Correlation Heatmap (FDR-BH)
Evrday_mem_err_tot_log (Day)
pTau217 (Time point)
1 2 3 4 5 6 7 8 9 Avg(9TP)
-14 0.47* 0.40 0.47* 0.53* 0.68** 0.51* 0.53* 0.42 0.39 0.52*
-13 0.43* 0.39 0.46* 0.50* 0.65** 0.47* 0.50* 0.40 0.41 0.50*
-12 0.41* 0.37 0.41* 0.47* 0.63** 0.44* 0.48* 0.38 0.37 0.47*
-11 0.32 0.28 0.36 0.37 0.52* 0.34 0.39 0.32 0.36 0.39
-10 0.31 0.28 0.34 0.37 0.53* 0.34 0.38 0.34 0.32 0.39
-9 0.37 0.35 0.39 0.46* 0.60** 0.42* 0.44* 0.37 0.34 0.44*
-8 0.30 0.26 0.32 0.37 0.55* 0.34 0.37 0.31 0.22 0.37
-7 0.38 0.36 0.40* 0.47* 0.63** 0.44* 0.45* 0.39 0.36 0.45*
-6 0.39 0.38 0.44* 0.46* 0.69** 0.44* 0.46* 0.56* 0.51* 0.46*
-5 0.40 0.42* 0.44* 0.50* 0.68** 0.45* 0.47* 0.52* 0.48* 0.47*
-4 0.37 0.35 0.38 0.46* 0.61** 0.43* 0.44* 0.37 0.34 0.43*
-3 0.37 0.35 0.37 0.46* 0.61** 0.43* 0.44* 0.35 0.33 0.43*
-2 0.36 0.33 0.37 0.44* 0.56* 0.40 0.43* 0.34 0.32 0.42*
-1 0.35 0.31 0.37 0.42 0.59* 0.40 0.42 0.38 0.38 0.41
1 (In-Lab) 0.33 0.30 0.35 0.40 0.50* 0.36 0.38 0.37 0.41 0.37
Pearson r
0.6
0.5
0.4
0.3

**Supplemental Figure 3.** Heatmaps showing the association between the p-Tau217 for each of the nine blood samples for backward cumulative averaging of the 15 study days for top panel - immediate recall, middle panel - delayed recall, and bottom panel - everyday memory errors. Correlation magnitudes shown are Pearson's r coefficient. Asterisks indicate significant associations that survive FDR correction and yellow crosses indicate the most significant associations.

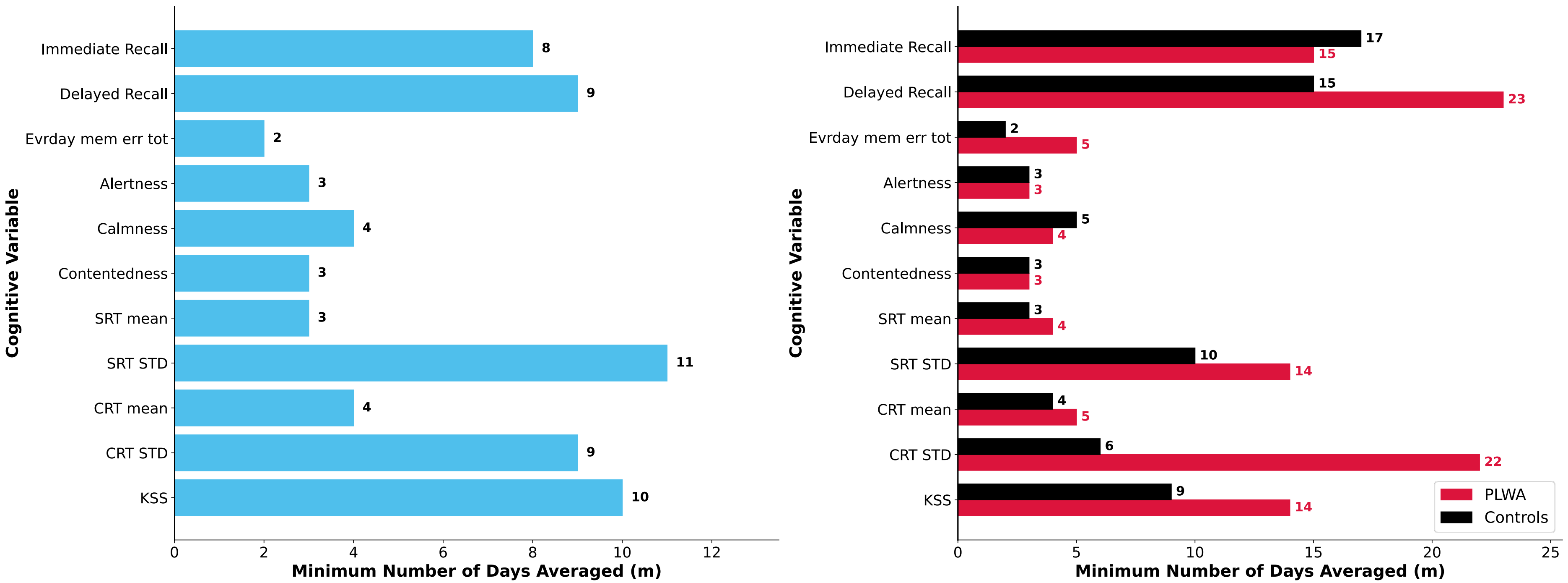


**Supplemental Figure 4.** Minimum number of days required (m) to achieve ICC > 0.9 for measures of daytime function for all participants (left panel) and separately for Controls (black) and PLWA (red) (right panel).
